# Evaluating the Potential of Wearable Technology in Early Stress Detection: A Multimodal Approach

**DOI:** 10.1101/2024.07.19.24310732

**Authors:** Basil A. Darwish, Nancy M. Salem, Ghada Kareem, Lamees N. Mahmoud, Ibrahim Sadek

## Abstract

Stress can adversely impact health, leading to issues like high blood pressure, heart diseases, and a compromised immune system. Consequently, using wearable devices to monitor stress is essential for prompt intervention and effective management. This study investigates the efficacy of wearable devices in the early detection of psychological stress, employing both binary and five-class classification models. Significant correlations were observed between stress levels and physiological signals, including Electrocardiogram (ECG), Electrodermal Activity (EDA), and Respiration (RESP), establishing these modalities as reliable biomarkers for stress detection. Utilizing the publicly available Wearable Stress and Affect Detection (WESAD) dataset, we employed two ensemble methods, Majority Voting (MV) and Weighted Averaging (WA), to integrate these signals, achieving maximum accuracies of 99.96% for binary classification and 99.59% for five-class classification. This integration significantly enhances the accuracy and robustness of the stress detection system. Furthermore, ten different classifiers were evaluated, and hyperparameter optimization and K-fold cross-validation ranging from 3-fold to 10-fold were applied. Both time-domain and frequency-domain features were examined separately. A review of commercially available wearable devices supporting these modalities was also conducted, resulting in recommendations for optimal configurations for practical applications. Our findings highlight the potential of multimodal wearable devices in advancing the early detection and continuous monitoring of psychological stress, with significant implications for future research and the development of improved stress detection systems.

## 1. Introduction

Stress, a condition of mental strain or pressure due to upsetting conditions, is a major contributor to human physiology and pathophysiology. It has been linked to several conditions, such as autoimmune diseases, metabolic syndrome, sleep disorders, and suicidal thoughts and inclination [1] Over 70% of Americans regularly experience stress. Chronic stress can severely impact physical, mental, and social behaviors, potentially leading to numerous serious human disorders [2]. It is associated with the development of cancer, cardiovascular disease, depression, and diabetes, and thus is deeply detrimental to physiological health and psychological wellbeing [3], [4].

The response to stressful stimuli is based on a complex brain network, which requires well-tuned, functional neuroanatomical processing to detect and interpret the event as a potential threat to humans. However, the difficulty with the diagnosis and treatment of stress disorders lies in the complexity of the system and in the fact that stressors trigger different structures of the brain network [1]. Traditional diagnostic methods, such as self-reported questionnaires and physiological measurements, often lack the accuracy and objectivity needed for effective stress management [5]. Furthermore, these methods provide spontaneous measures, and they are not best suited for long-term monitoring.

Research indicates stress significantly contributes to cardiovascular disease by triggering pathophysiological changes such as increased sympathetic activation, heightened blood pressure, and inflammatory responses. These stress-induced mechanisms are particularly critical in individuals with pre-existing cardiovascular conditions, emphasizing the need for targeted preventive measures [6].

Given the prevalence and impact of stress, developing robust methods for the rapid and accurate detection of human stress is of paramount importance. This is where Artificial Intelligence (AI) and Machine Learning (ML) come into play. AI and ML have been able to predict stress and detect the brain’s normal states vs. abnormal states (notably, in post-traumatic stress disorder (PTSD) with an accuracy around 90% [1], [7]. Recent advancements in ML techniques have further enhanced their predictive capabilities, making them valuable tools for stress detection [8].

Despite these advancements, several challenges and limitations exist in implementing ML for stress detection. Issues such as data privacy, the requirement for large and diverse datasets, and potential biases in algorithms need to be addressed [9]. Given that physiological signals have been demonstrated to be dependable indicators of stress [10], integrating multi-modal data (e.g., physiological signals, behavioral data, and environmental factors) could improve the robustness and accuracy of ML models [11].

The potential benefits of accurate stress detection using ML are extensive, including early intervention, personalized treatment plans, and improved mental health outcomes. To fully realize these benefits, future research should focus on refining ML algorithms, addressing ethical concerns, and developing user-friendly applications for both clinicians and patients [12]. In summary, while the application of ML in stress detection is promising, continuous research and development are crucial to overcome existing limitations and fully leverage these technologies to improve mental health care [13].

Given the critical importance of accurate stress detection and the potential of AI and ML technologies, it is essential to review the existing literature on these topics. The following section will provide a comprehensive examination of current research, highlighting key findings, methodological approaches, and the evolving landscape of AI and ML applications in stress detection. This review will also identify gaps in the literature and propose directions for future research, setting the stage for a deeper understanding of how these technologies can be harnessed to address the complexities of stress and its impact on human health.

## 2. Literature Review

Smets et al. (2016) [14] explores the application of machine learning techniques in detecting psychophysiological stress by analyzing various physiological signals. The research, conducted in a controlled laboratory setting, examines Electrocardiogram (ECG), Galvanic Skin Response (GSR), temperature, and respiration during a stress test. By comparing six different machine learning techniques, the study identifies personalized dynamic Bayesian networks as the most effective, achieving a prediction accuracy of 84.6%. The study’s approach of using both general and personal models to enhance classification accuracy is noteworthy. However, the research is confined to a controlled environment, which may not accurately represent real-world stress conditions. Furthermore, the study does not investigate the impact of stressor intensity or the potential for real-time stress monitoring outside the laboratory setting. Despite these limitations, the findings underscore the potential of machine learning in improving the precision of stress detection tools and contributing to the development of responsive stress management strategies.

Gjoreski et al. (2017) [15] explored a novel method for detecting stress using a wrist-worn device in real-life settings, combining machine learning with a context-based approach. This study employed multiple classifiers, including a Support Vector Machine (SVM) classifier, which achieved a 71% accuracy within a six-minute window. It tested three-class classification and utilized leave-one-subject-out (LOSO) cross-validation to ensure robustness. The research integrated data from a laboratory stress detector, an activity recognizer, and a context-based stress detector, aiming to provide continuous and unobtrusive stress monitoring that adapts to various activities and environmental factors. Despite its promising approach, the study acknowledges the challenges of accurately detecting stress in diverse real-life conditions and suggests the need for further refinement and broader testing.

Schmidt et al. (2018) [16] provide a significant contribution to affect recognition by offering a publicly available dataset specifically tailored for wearable stress detection. This multimodal dataset includes physiological and motion data collected from both wrist- and chest-worn devices during a controlled lab study involving 15 subjects. Key modalities encompassed include blood volume pulse (BVP), electrocardiogram (ECG), electrodermal activity (EDA), electromyogram (EMG), respiration (RESP), body temperature (TEMP), and three-axis acceleration (ACCE). Notably, the dataset was designed to bridge gaps in available standard datasets by including multiple affective states, such as neutral, stress, and amusement, and supports the development of automated stress monitoring systems. To validate their approach, the researchers employed the LOSO cross-validation method. They report a classification accuracy of 93.12% using a linear discriminant analysis classifier (LDA) for binary stress detection. This high level of accuracy highlights the robustness of the dataset and the effectiveness of the LDA classifier in stress and affect detection. However, the study does present certain limitations. It does not explore the impact of varying window sizes for data analysis, which could affect the performance and applicability of the classification algorithms in real-world scenarios where stress levels can fluctuate over different periods.

Can et al. (2019) [17] developed a portable stress detection system that utilizes physiological data gathered from discreet smart wearables. The system incorporates techniques for artifact removal and feature extraction tailored for real-world applications. They gathered physiological signals and questionnaire data from 21 participants using Samsung Gear S, S2, and Empatica E4 sensors. The analysis, employing a 10-fold cross-validation for robustness, revealed that the Multilayer Perceptron (MLP) algorithm yielded the highest accuracy of 92.19% using heart rate (HR) and ACCE data from the Empatica E4. However, a limitation of this study is its focus on binary stress classification, rather than employing a multi-class classification approach that could potentially offer a more nuanced understanding of stress levels.

Siirtola et al. (2020) [18] utilizes the publicly available AffectiveROAD dataset, which includes data collected via the Empatica E4 sensor. In their experiments, the integration of features from blood volume pulse (BVP) and skin temperature (SKT) yielded the most favorable results. Using LOSO cross-validation, the study achieved an average accuracy of 82.3% with a bagged tree-based ensemble. However, a notable limitation of the study is the absence of class-balancing techniques, which could potentially affect the reliability and generalizability of the findings.

Kaczor et al. (2020) [19] address physician stress in emergency medicine through the objective monitoring of physiological indicators and digital biomarkers using wearable sensors. Their innovative approach deviates from traditional subjective self-reports by employing machine learning algorithms, to determine stress episodes. The study achieves a prediction accuracy of 64.5% using a Naive Bayes classifier within a 20-minute pre-stress episode window. Notably, the research employs robust validation techniques, including 10-fold cross-validation and Receiver Operating Characteristic (ROC) curve analysis, and tests multiple classifiers to ensure reliability. However, the study is limited by its exclusive use of binary classification, failing to account for varying stress levels. It does not explore the effects of different time window sizes on prediction accuracy. Despite these limitations, the findings highlight the potential of wearable sensors in facilitating real-time stress interventions and improving the well-being of high-stress professionals.

Iqbal et al. (2021) [20] investigated the utility of wearable sensors for the real-time monitoring of stress. The study utilized the WESAD dataset. The study applied logistic regression to assess features derived from electromyography (EMG), electrodermal activity (EDA), respiration (RESP), and heart rate (HR). The logistic regression model achieved an accuracy of 85.71% in binary classification of stress states. This high level of accuracy demonstrates the potential of wearable devices in effectively identifying stress. Additionally, the validity of the approach was confirmed through 14-fold cross-validation, underlining the efficacy of logistic regression in processing complex bio physiological data for stress detection.

Iqbal et al. (2022) [21] delve into advanced strategies for continuous and real-time stress monitoring. Their research emphasizes the use of wearable sensor technology, with a particular focus on heart rate (HR) as a vital physiological parameter. Employing a Random Forest classifier integrated with 10-fold cross-validation, they analyze the SWELL-KW dataset. This technique not only enhances the robustness of their findings but also facilitates a significant binary classification of stress, achieving an accuracy of 75%.

Ehrhart et al. (2022) [22] highlight the significant advancements in human-centered applications leveraging wearable sensors and machine learning, particularly deep learning, for stress detection through physiological signals. They specifically utilized a stacked Long Short-Term Memory (LSTM) and a Fully Convolutional Network (FCN) classifier, employing features based on Galvanic Skin Response (GSR) and Skin Temperature (SKT), to achieve an impressive binary classification accuracy of 86.76% using tests on unseen cross-validation. The authors discuss the challenges of acquiring large, labeled datasets in this domain, which often leads to imbalanced data for training robust models. To address these limitations, they explored the use of a Conditional Generative Adversarial Network (cGAN) to augment the dataset, effectively enhancing its volume and diversity. This approach not only mitigated data imbalance but also significantly improved classifier performance, demonstrating that the synthetic data are indistinguishable from real data in their application. Despite these advancements, the study is limited by the absence of multi-class classification capabilities, which could potentially enhance the applicability of the stress detection models further.

Kuttala et al. (2023) [23] conducted a pivotal study on binary stress classification utilizing advanced deep learning techniques. The research spotlighted a critical issue: individuals experiencing stress often fail to recognize their stress levels, thus underlining the necessity for early and precise stress detection mechanisms. In their innovative approach, they harnessed multimodal hierarchical CNN feature fusion, significantly enhancing stress detection capabilities. This technique involved the integration of low, mid, and high-level features from Convolutional Neural Networks (CNNs), with concatenated multi-level CNN features for each of two key physiological signals: Electrodermal Activity (EDA) and Electrocardiogram (ECG). These features were then synergistically fused using the Multimodal Transfer Module (MMTM). Their comprehensive analysis spanned both raw frequency domain data and targeted frequency band features to ascertain the model’s effectiveness. The empirical testing of the model across four benchmark datasets—ASCERTAIN, CLAS, MAUS, and WAUC—involved an initial training phase with 36, 18, 43, and 42 subject samples, respectively, followed by a testing phase comprising 9, 4, 16, and 16 subject samples from each dataset. The results were impressive, demonstrating high accuracies of 97.61% on ASCERTAIN, 95.94% on CLAS, 88.75% on MAUS, and 83.96% on WAUC. Despite these promising outcomes, the study acknowledged the need to expand the studies on hierarchical feature fusion and different multi-modal fusion techniques on hierarchical features.

Kalra et al. (2023) [24] conducted a study on pulse rate variability (PRV) using photoplethysmography (PPG) to monitor 15 subjects across five cognitive states: relaxation, deep breathing, and three varying levels of stress-related tasks. They discovered 18 significant features, split evenly between the time and frequency domains, which all showed statistical significance (p < 0.05) according to the Friedman test. Initially, a multi-layer perceptron (MLP) model was employed, achieving a classification accuracy of 85.1%±1.1%. This was further improved to 91%±1.1% when deep neural networks (DNN) were applied. A potential limitation is its broader focus on multiple cognitive states rather than exclusively on stress. This may dilute the specific insights and nuances related to stress detection and analysis, potentially impacting the specificity of the findings related to stress-related pulse rate variability.

Greco et al. (2023) [25] developed a novel methodology for detecting acute stress using only electrodermal activity (EDA) signals. This approach utilized a Support Vector Machine with a Recursive Feature Elimination algorithm (SVM-RFE) for classifying stress at an individual level. Employing a single-sensor system, the method demonstrated robustness to noise, incorporated rigorous phasic decomposition, and implemented unbiased multiclass classification. The methodology was tested on 65 volunteers subjected to various acute stress stimuli through a modified Trier Social Stress Test. For binary classification, the authors reported successful stress detection with an average accuracy of 94.62%. Furthermore, they proposed a four-class pattern recognition system capable of distinguishing between non-stressed states and three different stress conditions, achieving an average accuracy of 75% using leave-one-subject-out (LOSO) cross-validation. These results, obtained under controlled conditions, lay the groundwork for future applications in more ecological settings.

Richer et al. (2024) [26] explored the association between acute psychosocial stress and body movements using inertial measurement unit (IMU)-based motion capture suits. Data were gathered from 59 participants over two studies, in which participants experienced both the Trier Social Stress Test (TSST) and a control condition (friendly-TSST; f-TSST) in a randomized sequence. The research revealed a consistent freezing behavior in response to acute stress, marked by decreased overall movement and longer periods of immobility. Utilizing a Random Forest (RF) classifier with five-fold cross-validation, the study achieved a 73.4% accuracy in identifying acute stress from movement data. However, the study was limited by its binary classification approach, which did not address multiple stress levels. This study demonstrates the potential of using body posture and movement analysis as reliable indicators of acute psychosocial stress, presenting an alternative to conventional stress assessment methods.

In light of the gaps identified in the current literature, this paper aims to investigate the validity of using wearable devices for the early detection of psychological stress in both binary and five-class classifications. We employ machine learning techniques, testing various classifiers with optimized hyperparameters and cross-validation, using ECG, EDA, and RESP biometrics individually and in ensemble. Our study uniquely contributes by exploring five-class stress classification and an ensemble system using multiple biometrics, aiming to improve quality of life through more reliable and nuanced stress monitoring.

## 3. Methodology

To address the research gaps identified earlier, this study aims to explore the feasibility of robust psychological stress detection employing systematic signal segmentation and feature extraction to comprehensively characterize physiological responses, a rigorous machine learning pipeline involving feature selection, diverse classifiers, and hyperparameter optimization, a meticulous performance evaluation utilizing K-fold cross-validation (K-CV) as well as binary and multi-class detection. A comprehensive overview of our workflow is presented in Fig. 1.

**Fig. 1:**
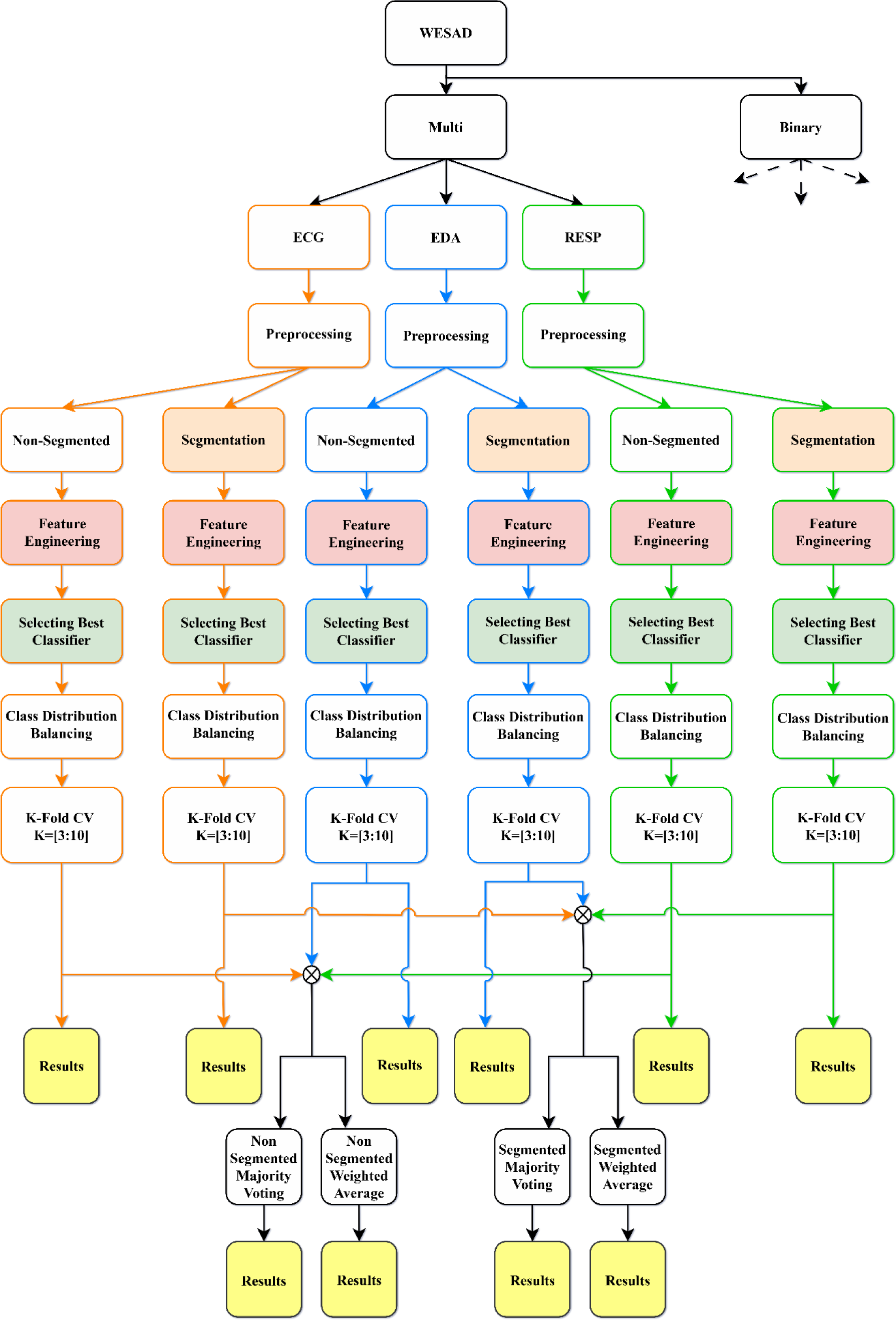
Comprehensive binary and multi-class stress classification workflow for preprocessing, segmentation, feature engineering, and classifier selection with hyperparameter optimization of ECG, EDA, and RESP modalities from the WESAD dataset.

### 3.1. Data Acquisition

This study employs the Wearable Stress and Affect Detection (WESAD) dataset, a publicly available resource for stress classification research (https://ubicomp.eti.uni-siegen.de/home/datasets/icmi18/) [16]. Within WESAD, data for 15 subjects was collected across three experimental conditions designed to elicit varying stress levels: baseline, stress, and amusement. This study utilizes an electrocardiogram (ECG), electrodermal activity (EDA), and respiration (RESP) collected from the chest-worn RespiBAN Professional device, all sampled at 700 Hz. The labels used in this study were from the Positive and Negative Affect Schedule questionnaire (PANAS) available in WESAD, more specifically the 21st item (Stressed) with its five possible responses (1 = Not at all, 2 = A little bit, 3 = Somewhat, 4 = Very much, 5 = Extremely) in case of multi-class. For binary classification, responses of ‘Not at all’ were considered class 0 (no stress) with remaining response levels considered class 1 (stressed).

### 3.2. Pre-processing

We utilized the BioSPPy library, an open-source tool for biosignal processing. This library provided us with robust and efficient algorithms for the analysis and filtering of Electrocardiogram (ECG), Electrodermal Activity (EDA), and Respiration (RESP) signals. For this study, we used the default parameters provided by the library. For additional information, documentation, and code examples, we recommend visiting the official BioSPPy GitHub repository (https://github.com/PIA-Group/BioSPPy).

To thoroughly investigate the impact of temporal signal length on stress classification, this study employed a strategic segmentation approach inspired by previous research [16], [27]. Window sizes were tested in increments, exploring durations of 60, 120, 210, 300, and 390 seconds. Additionally, to examine the effect of overlap, shifts of 10, 20, 30, 60, 120, 210, 300, and 390 seconds were applied to each window size appropriately to a total of 31 combinations, including the original unsegmented signal.

To ensure feature compatibility and improve machine learning model performance, the extracted features were normalized using Z-score. This process involved subtracting the mean and dividing by the standard deviation of each feature.

### 3.3. Feature Extraction

Feature extraction targeted both time-domain and frequency-domain characteristics across modalities. The statistical results were obtained directly from preprocessed signal segments. To analyze frequency patterns, a Fast Fourier Transform (FFT) and power spectral density (PSD) calculations were performed. The same set of statistical features was then derived from the power spectrum to enable comparative analysis across domains (Table 1)

**Table 1:** Extracted Features, including time and frequency domain features.

| Modality | Time Domain Features | Frequency Domain Features |
| --- | --- | --- |
| ECG/ EDA/<br>RESP | Mean | PSD Mean |
|  | Variance | PSD Variance |
|  | Standard Deviation | PSD Standard Deviation |
|  | Median | PSD Median |
|  | Maximum | PSD Maximum |
|  | Minimum | PSD Minimum |
|  | First Quartile | PSD First Quartile |
|  | Third Quartile | PSD Third Quartile |
|  | Skewness | PSD Skewness |
|  | Kurtosis | PSD Kurtosis |

### 3.4. Feature Selection

In our study, feature selection was performed using the Select From Model (SFM) method [28], which employs a Random Forest classifier as a meta-transformer. Specifically, we utilized a Random Forest with n=100 trees to determine feature importance scores, following a methodology analogous to that described in [26].

### 3.5. Classifiers and Hyperparameter Optimization

To ensure a comprehensive evaluation and the potential to uncover unexpected relationships; we tested ten different classifiers: Random Forest (RF), Extreme Gradient Boosting (XGB), k-nearest Neighbors (kNN), Logistic Regression (LR), Decision Tree (DT), AdaBoost (AB), Extra Trees (ET), Bagging (BAG), Quadratic Discriminant Analysis (QDA), and Linear Discriminant Analysis (LDA). We also applied hyperparameter optimization using grid search.

### 3.6. Class Distribution Balancing

Addressing class imbalance within the dataset was essential in mitigating potential classifier bias, Generative Adversarial Networks (GANs) have demonstrated effectiveness in creating novel data samples, making them a valuable technique for augmenting datasets. This augmentation is particularly beneficial for enhancing classifier performance on datasets that are small or imbalanced [22], [29]. Hence, we employed a multi-step Generative Adversarial Network (GAN) based data augmentation strategy. First, class representation was calculated, and classes exceeding the mean representation were truncated through random subsampling. Under-represented classes were targeted for augmentation with dedicated GANs [30]. These GANs were trained to learn underlying feature distributions, enabling the generation of synthetic samples mimicking the real data’s characteristics. Augmentation continued until each under-represented class matched the mean class frequency, creating a more balanced dataset for training.

### 3.7. Evaluation

The best-performing optimized model for each modality was utilized. To ensure a robust evaluation and mitigate potential performance variance due to data splits, K-fold cross-validation (K-CV) with K values ranging from 3 to 10 was employed [31]. This approach provides a more reliable estimate of performance compared to a single train/test split, as it reduces the risk of overfitting and helps assess the model’s generalization to unseen data. Ensemble methods, specifically majority voting (MV) and weighted averaging (WA), were applied to the outputs of ECG, EDA, and RESP. This investigation aimed to determine potential performance gains from a multi-modal approach compared to evaluating each modality independently. Accuracy (ACC) quantifies the proportion of correct predictions, while Precision (P) measures the accuracy of identifying positive labels correctly. Recall (R) indicates the percentage of actual positive cases the model successfully identifies. The F-measure (F1) score, the harmonic mean of precision and recall, provides a single metric balancing these two aspects. Additionally, the Area Under the Receiver Operating Characteristic Curve (AUC) was used to evaluate the model’s performance, reflecting its ability to distinguish between positive and negative classes. These five evaluation metrics were calculated for each fold and averaged across K-CV runs, offering a comprehensive assessment of model performance. The equations are defined below.

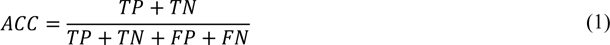

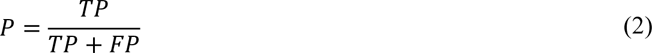

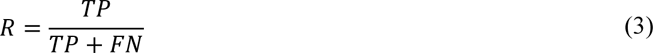

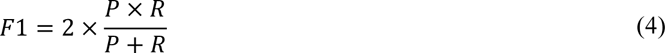

## 4. Results

This study evaluated the efficacy of time-domain and frequency-domain statistical features including common features such as mean, variance, and others extracted from ECG, EDA, and RESP for the early detection of psychological stress (binary and multi-class classification). Ten different machine learning (ML) models were tested across these modalities to identify the most effective configurations for stress detection. K-fold cross-validation (K-CV) with K ranging from 3 to 10 was employed to enhance the generalizability and robustness of the results by evaluating performance on multiple non-overlapping data splits. To further leverage the information contained in each modality, we explored the performance of two ensemble methods: MV and WA. These methods combine predictions from individual modalities to potentially achieve improved classification.

### 4.1. Time-domain

Ensemble methods demonstrably outperformed individual modalities across various configurations for both binary and multiclass classification, as visually depicted in Fig. 2 and Fig. 3 respectively. Fig. 4 depicts the ROC curve for non-overlap WA average binary classification. Table 2 provides a consolidated overview of the peak performance achieved for each modality, classification type, and ensemble method.

**Fig. 2:**
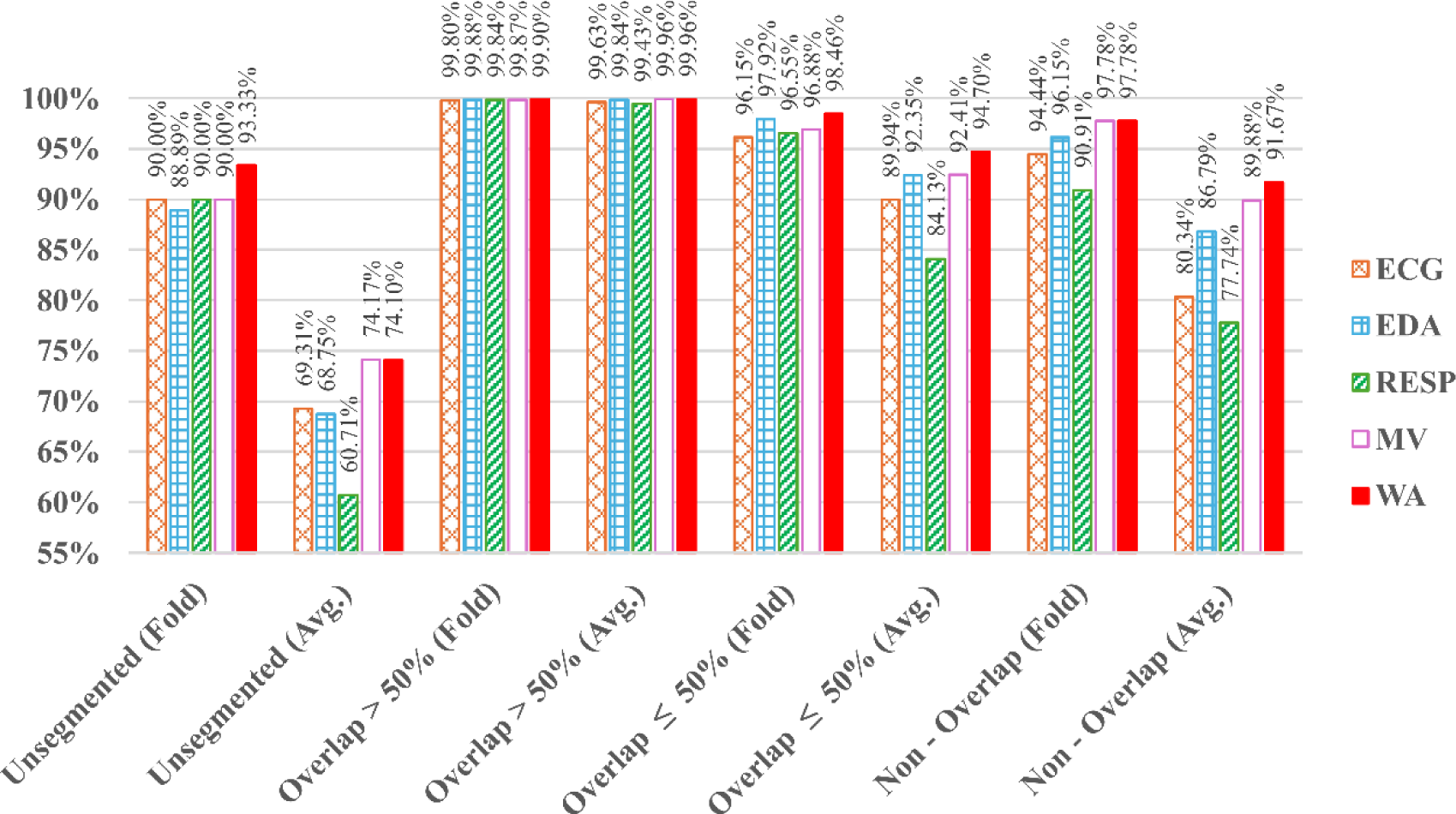
Best binary classification results based on time domain features.

**Fig. 3:**
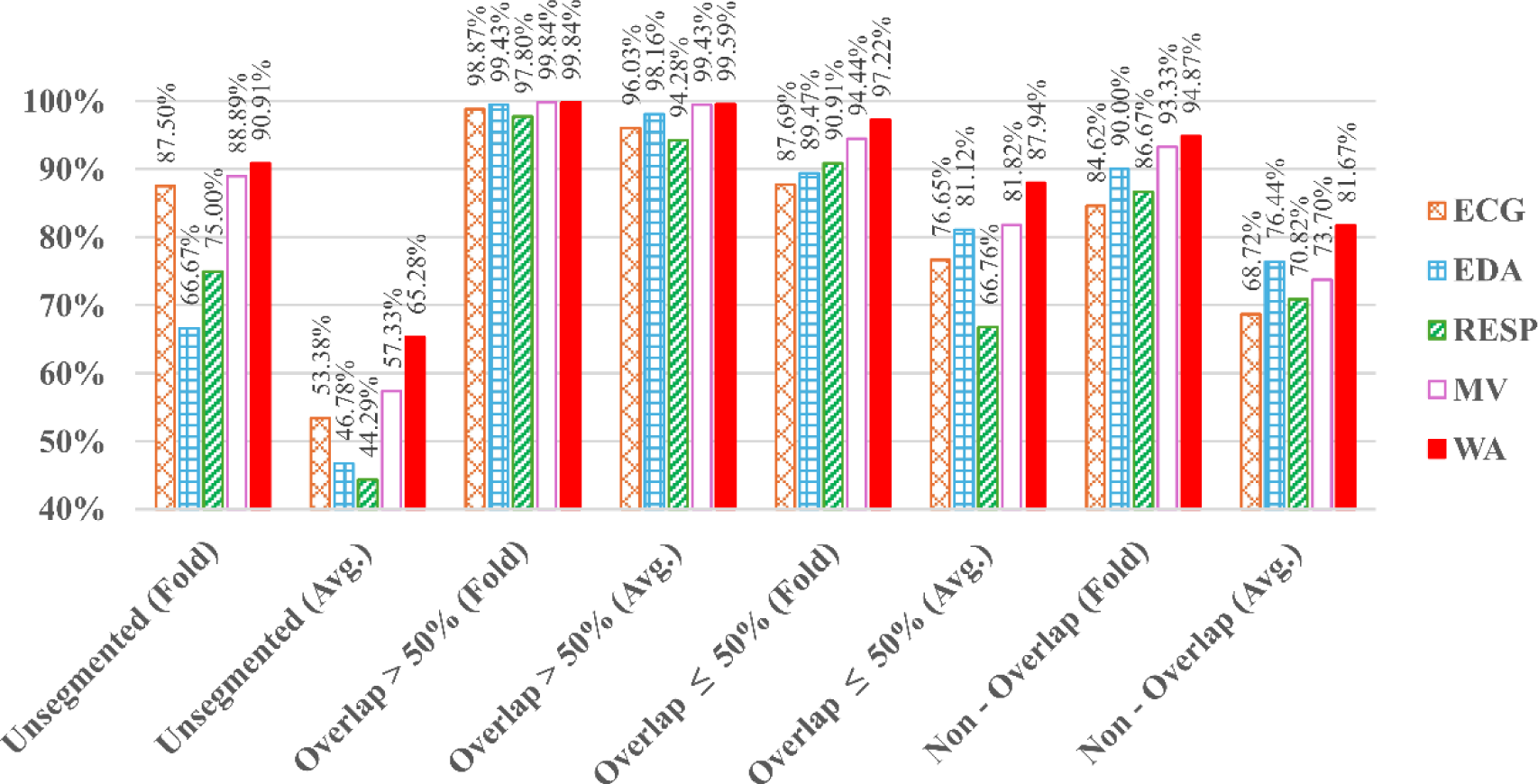
Best multi-class classification results based on time domain features.

**Fig. 4:**
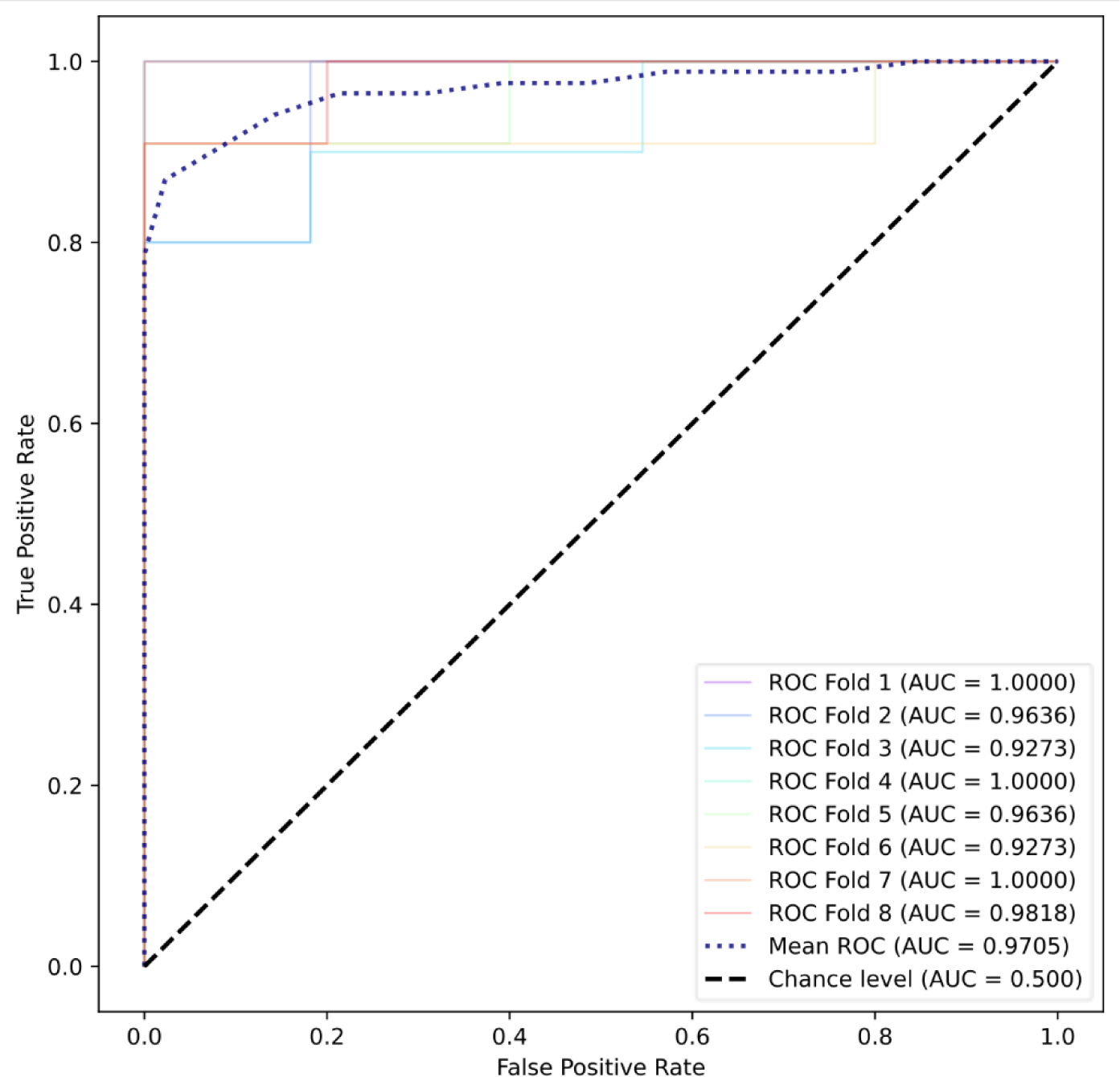
Binary classification WA 210_210 ROC curve based on time domain features.

**Table 2:**
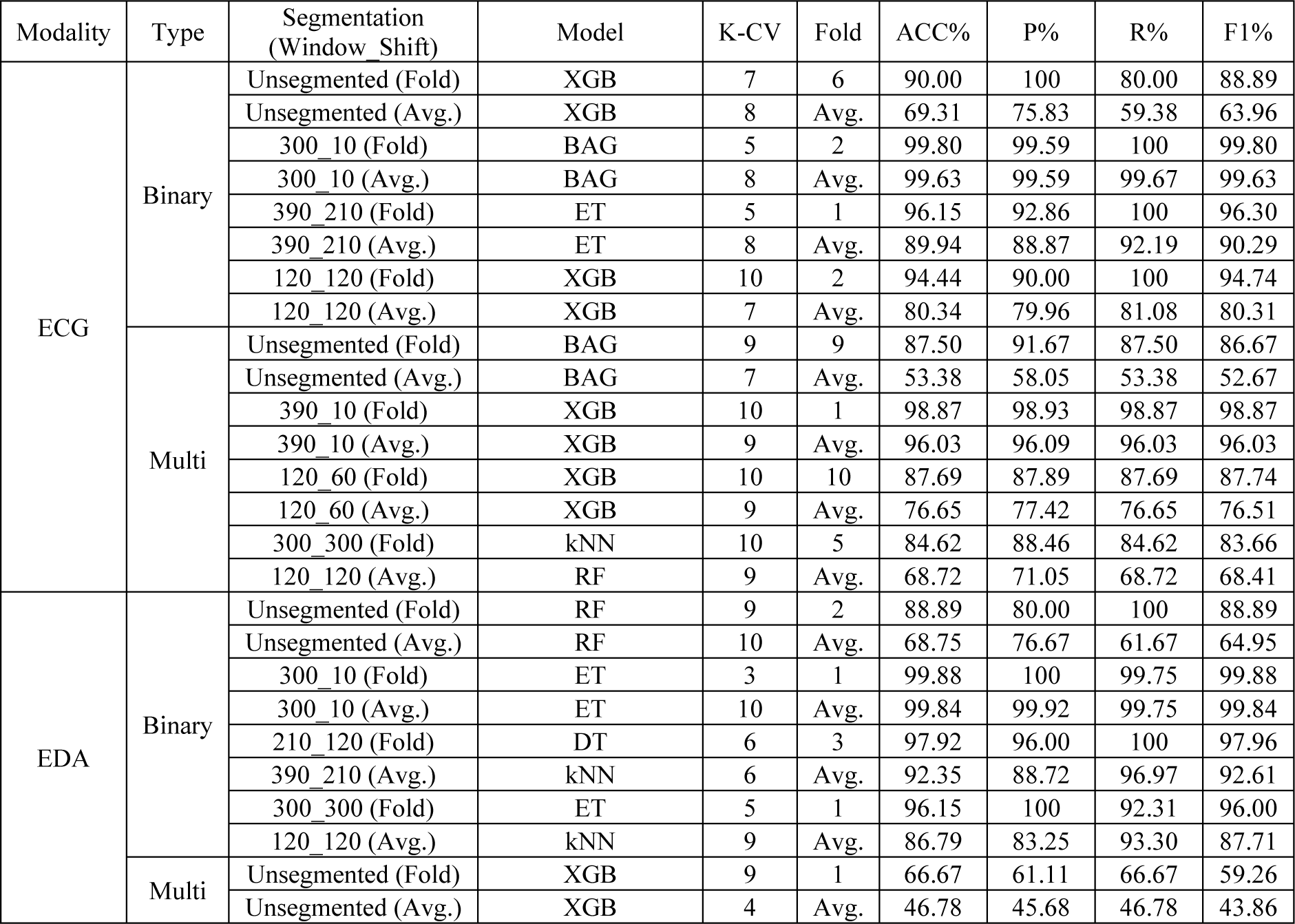
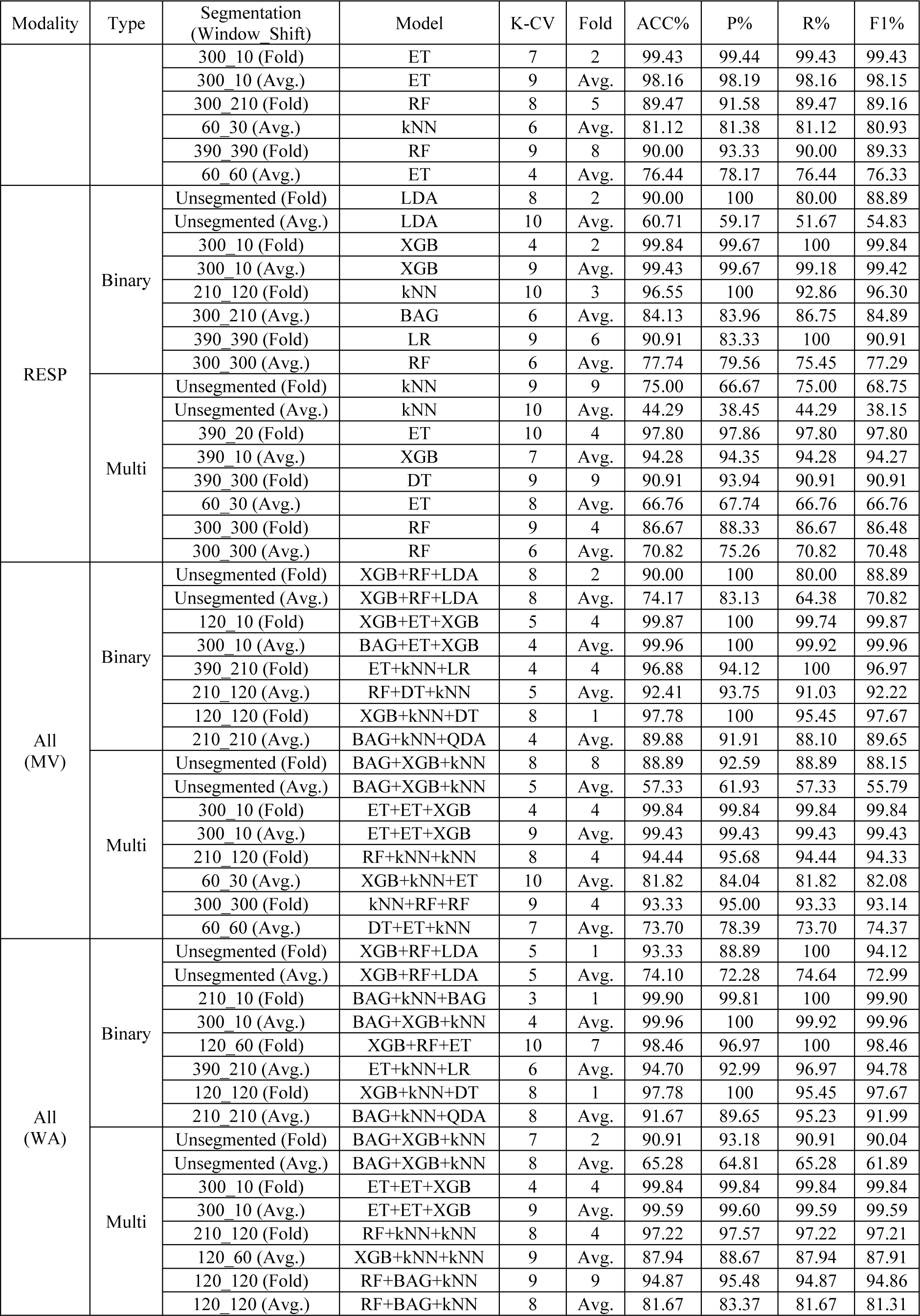
Best results based on time domain features.

#### 4.1.1 Performance Evaluation of ECG Modality

As presented in Table 2, the performance of the ECG modality was assessed across different segmentation strategies and classifiers for binary classification. With unsegmented ECG data, the optimized XGB classifier achieved the highest accuracy of 69.31% using 8-CV. For segmentations with over 50% overlap, a window of 300 seconds with a 10-second shift yielded the highest accuracy of 99.63% using an optimized BAG classifier and 8-CV. In cases of segmentation with 50% and under overlap, a window of 390 seconds with a 210-second shift resulted in the highest accuracy of 89.94% with an optimized ET classifier and 8-CV. For segmentation with no overlap, a window of 120 seconds provided the highest accuracy of 80.34% using an optimized XGB classifier and 7-CV. These results are illustrated in Fig. 2, providing a visual summary of the performance across different segmentation strategies.

As detailed in Table 2, the evaluation of the ECG modality for multi-class classification revealed varying performance across different segmentation approaches and classifiers. With unsegmented ECG data, the optimized BAG classifier achieved the highest accuracy of 53.38% using 7-CV. For segmentations with over 50% overlap, a window of 390 seconds with a 10-second shift yielded the highest accuracy of 96.03% using an optimized XGB classifier and 9-CV. In cases of segmentation with 50% and under overlap, a window of 120 seconds with a 60-second shift resulted in the highest accuracy of 76.65% with an optimized XGB classifier and 9-CV. For segmentation with no overlap, a window of 120 seconds provided the highest accuracy of 68.72% using an optimized RF classifier and 9-CV. Fig. 3 visually summarizes these results, highlighting the performance variations across different segmentation strategies.

#### 4.1.2 Performance Evaluation of EDA Modality

As presented in Table 2, the performance of the EDA modality was assessed across different segmentation strategies and classifiers for binary classification. With unsegmented EDA data, the optimized RF classifier achieved the highest accuracy of 68.75% using 10-CV. For segmentations with over 50% overlap, a window of 300 seconds with a 10-second shift yielded the highest accuracy of 99.84% using an optimized ET classifier and 10-CV. In cases of segmentation with 50% and under overlap, a window of 390 seconds with a 210-second shift resulted in the highest accuracy of 92.35% with an optimized kNN classifier and 6-CV. For segmentation with no overlap, a window of 120 seconds provided the highest accuracy of 86.79% using an optimized kNN classifier and 6-CV. These results are illustrated in Fig. 2, providing a visual summary of the performance across different segmentation strategies.

As detailed in Table 2, the evaluation of the EDA modality for multi-class classification revealed varying performance across different segmentation approaches and classifiers. With unsegmented EDA data, the optimized XGB classifier achieved the highest accuracy of 46.78% using 4-CV. For segmentations with over 50% overlap, a window of 300 seconds with a 10-second shift yielded the highest accuracy of 98.16% using an optimized ET classifier and 9-CV. In cases of segmentation with 50% and under overlap, a window of 60 seconds with a 30-second shift resulted in the highest accuracy of 81.12% with an optimized kNN classifier and 6-CV. For segmentation with no overlap, a window of 60 seconds provided the highest accuracy of 76.44% using an optimized ET classifier and 4-CV. Fig. 3 visually summarizes these results, highlighting the performance variations across different segmentation strategies.

#### 4.1.3 Performance Evaluation of RESP Modality

As presented in Table 2, the performance of the RESP modality was assessed across different segmentation strategies and classifiers for binary classification. With unsegmented RESP data, the optimized LDA classifier achieved the highest accuracy of 60.71% using 10-CV. For segmentations with over 50% overlap, a window of 300 seconds with a 10-second shift yielded the highest accuracy of 99.43% using an optimized XGB classifier and 9-CV. In cases of segmentation with 50% and under overlap, a window of 300 seconds with a 210-second shift resulted in the highest accuracy of 84.13% with an optimized BAG classifier and 6-CV. For segmentation with no overlap, a window of 300 seconds provided the highest accuracy of 77.74% using an optimized RF classifier and 6-CV. These results are illustrated in Fig. 2, providing a visual summary of the performance across different segmentation strategies.

As detailed in Table 2, the evaluation of the RESP modality for multi-class classification revealed varying performance across different segmentation approaches and classifiers. With unsegmented RESP data, the optimized kNN classifier achieved the highest accuracy of 44.29% using 10-CV. For segmentations with over 50% overlap, a window of 390 seconds with a 10-second shift yielded the highest accuracy of 94.28% using an optimized XGB classifier and 7-CV. In cases of segmentation with 50% and under overlap, a window of 60 seconds with a 30-second shift resulted in the highest accuracy of 66.76% with an optimized ET classifier and 8-CV. For segmentation with no overlap, a window of 300 seconds provided the highest accuracy of 70.82% using an optimized RF classifier and 6-CV. Fig. 3 visually summarizes these results, highlighting the performance variations across different segmentation strategies.

#### 4.1.4 Performance Evaluation of Modalities MV Ensemble Method

As presented in Table 2, the performance of the modalities MV ensemble was assessed across different segmentation strategies and classifiers for binary classification. With unsegmented MV Ensemble data, the optimized XGB, RF, and LDA classifiers for ECG, EDA, and RESP respectively achieved the highest accuracy of 74.17% using 8-CV. For segmentations with over 50% overlap, a window of 300 seconds with a 10-second shift yielded the highest accuracy of 99.96% using optimized BAG, ET, and XGB classifiers for ECG, EDA, and RESP respectively, and 4-CV. In cases of segmentation with 50% and under overlap, a window of 210 seconds with a 120-second shift resulted in the highest accuracy of 92.41% with optimized RF, DT, and kNN classifiers for ECG, EDA, and RESP respectively, and 5-CV. For segmentation with no overlap, a window of 210 seconds provided the highest accuracy of 89.88% using optimized BAG, kNN, and QDA classifiers for ECG, EDA, and RESP respectively, and 4-CV. These results are illustrated in Fig. 2, providing a visual summary of the performance across different segmentation strategies.

As detailed in Table 2, the evaluation of the modalities MV ensemble for multi-class classification revealed varying performance across different segmentation approaches and classifiers. With unsegmented MV Ensemble data, the optimized BAG, XGB, and kNN classifiers for ECG, EDA, and RESP respectively achieved the highest accuracy of 57.33% using 5-CV. For segmentations with over 50% overlap, a window of 300 seconds with a 10-second shift yielded the highest accuracy of 99.43% using optimized ET, ET, and XGB classifiers for ECG, EDA, and RESP respectively, and 9-CV. In cases of segmentation with 50% and under overlap, a window of 60 seconds with a 30-second shift resulted in the highest accuracy of 81.82% with optimized XGB, kNN, ET classifiers for ECG, EDA, and RESP respectively, and 10-CV. For segmentation with no overlap, a window of 60 seconds provided the highest accuracy of 73.70% using optimized XGB, ET, and kNN classifiers for ECG, EDA, and RESP respectively, and 7-CV. Fig. 3 visually summarizes these results, highlighting the performance variations across different segmentation strategies.

#### 4.1.5 Performance Evaluation of Modalities WA Ensemble Method

As presented in Table 2, the performance of the modalities WA ensemble was assessed across different segmentation strategies and classifiers for binary classification. With unsegmented WA Ensemble data, the optimized XGB, RF, and LDA classifiers for ECG, EDA, and RESP respectively achieved the highest accuracy of 74.10% using 5-CV. For segmentations with over 50% overlap, a window of 300 seconds with a 10-second shift yielded the highest accuracy of 99.96% using optimized BAG, XGB, and kNN classifiers for ECG, EDA, and RESP respectively, and 4-CV. In cases of segmentation with 50% and under overlap, a window of 390 seconds with a 210-second shift resulted in the highest accuracy of 94.70% with optimized ET, kNN, LR classifiers for ECG, EDA, and RESP respectively, and 6-CV. For segmentation with no overlap, a window of 210 seconds provided the highest accuracy of 91.67% using optimized BAG, kNN, and QDA classifiers for ECG, EDA, and RESP respectively and 8-CV, additionally ROC is depicted in Fig. 4. These results are illustrated in Fig. 2, providing a visual summary of the performance across different segmentation strategies.

As detailed in Table 2, the evaluation of the modalities WA ensemble for multi-class classification revealed varying performance across different segmentation approaches and classifiers. With unsegmented WA Ensemble data, the optimized BAG, XGB, and kNN classifiers for ECG, EDA, and RESP respectively achieved the highest accuracy of 65.28% using 8-CV. For segmentations with over 50% overlap, a window of 300 seconds with a 10-second shift yielded the highest accuracy of 99.59% using optimized ET, ET, and XGB classifiers for ECG, EDA, and RESP respectively, and 9-CV. In cases of segmentation with 50% and under overlap, a window of 120 seconds with a 60-second shift resulted in the highest accuracy of 87.94% with optimized XGB, kNN, kNN classifiers for ECG, EDA, and RESP respectively, and 9-CV. For segmentation with no overlap, a window of 120 seconds provided the highest accuracy of 81.67% using optimized RF, BAG, and kNN classifiers for ECG, EDA, and RESP respectively, and 8-CV. Fig. 3 visually summarizes these results, highlighting the performance variations across different segmentation strategies.

### 4.2. Frequency-domain

Ensemble methods mostly maintained their dominance in the frequency domain for both binary and multiclass classification, as depicted in Fig. 5 and Fig. 6 respectively. Fig. 7 depicts the ROC curve for non-overlap WA average binary classification. Table 3 summarizes the peak performance achieved for each modality, classification type, and ensemble method.

**Fig. 5:**
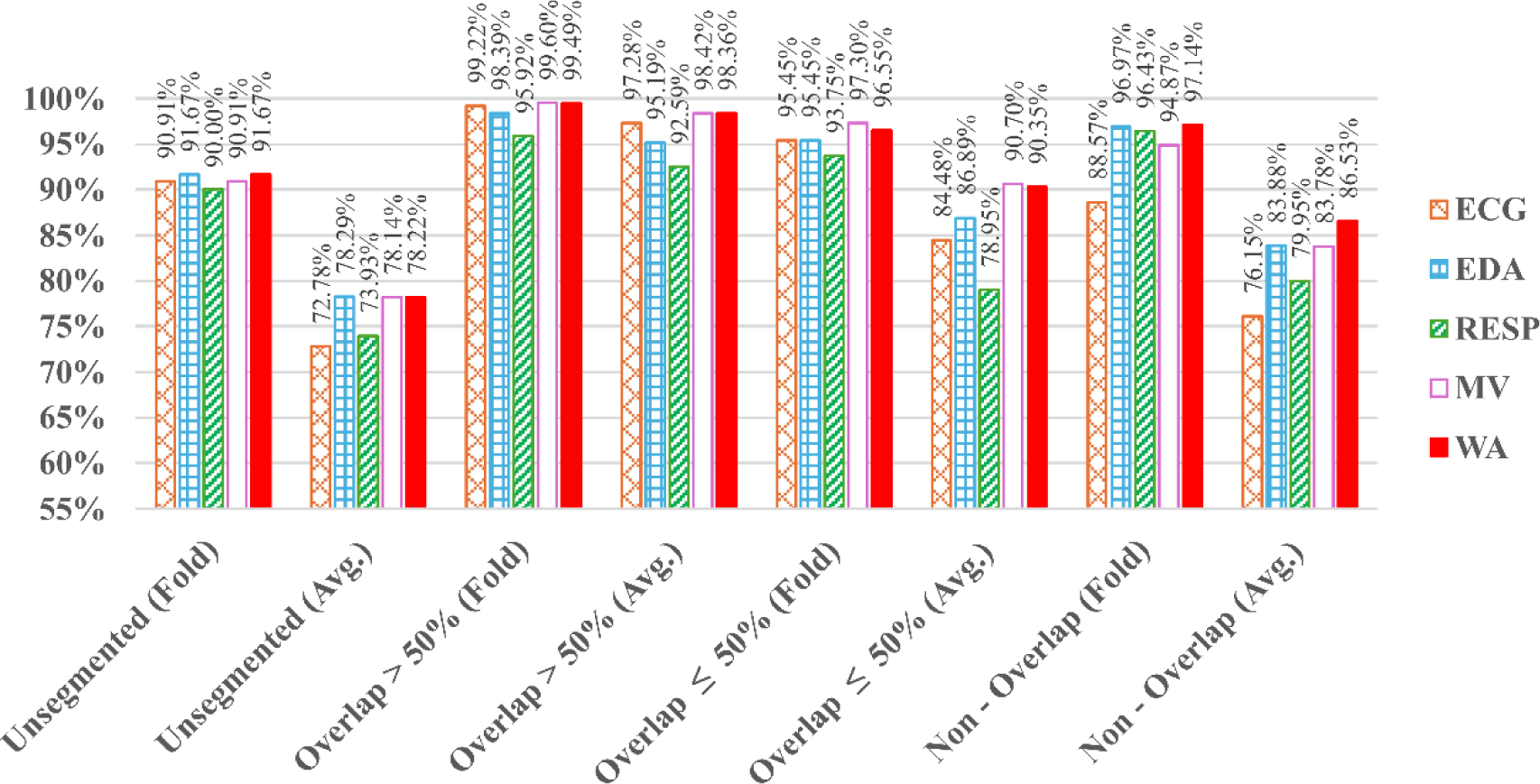
Best binary classification results based on frequency domain features.

**Fig. 6:**
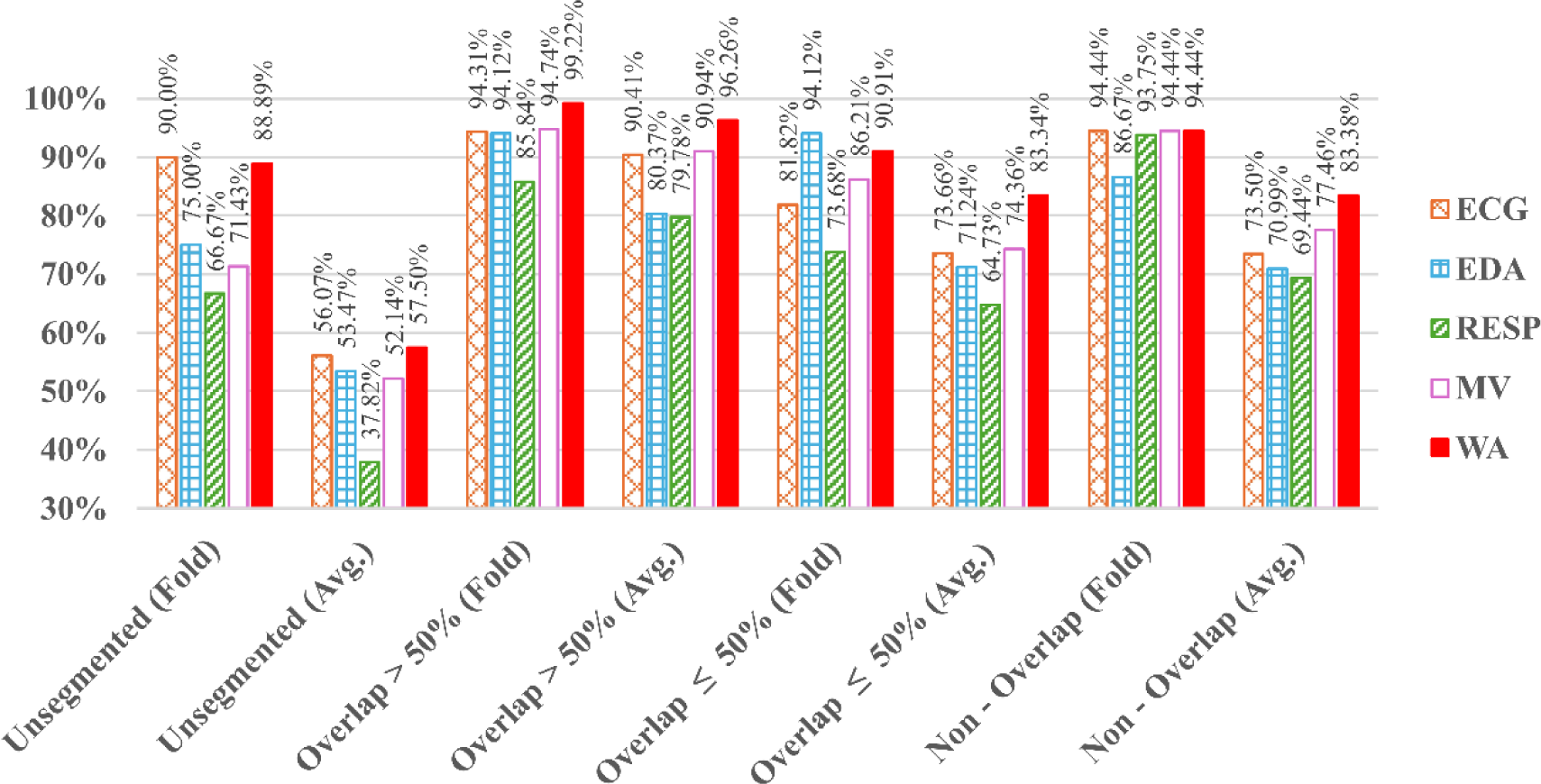
Best multi-class classification results based on frequency domain features.

**Fig. 7:**
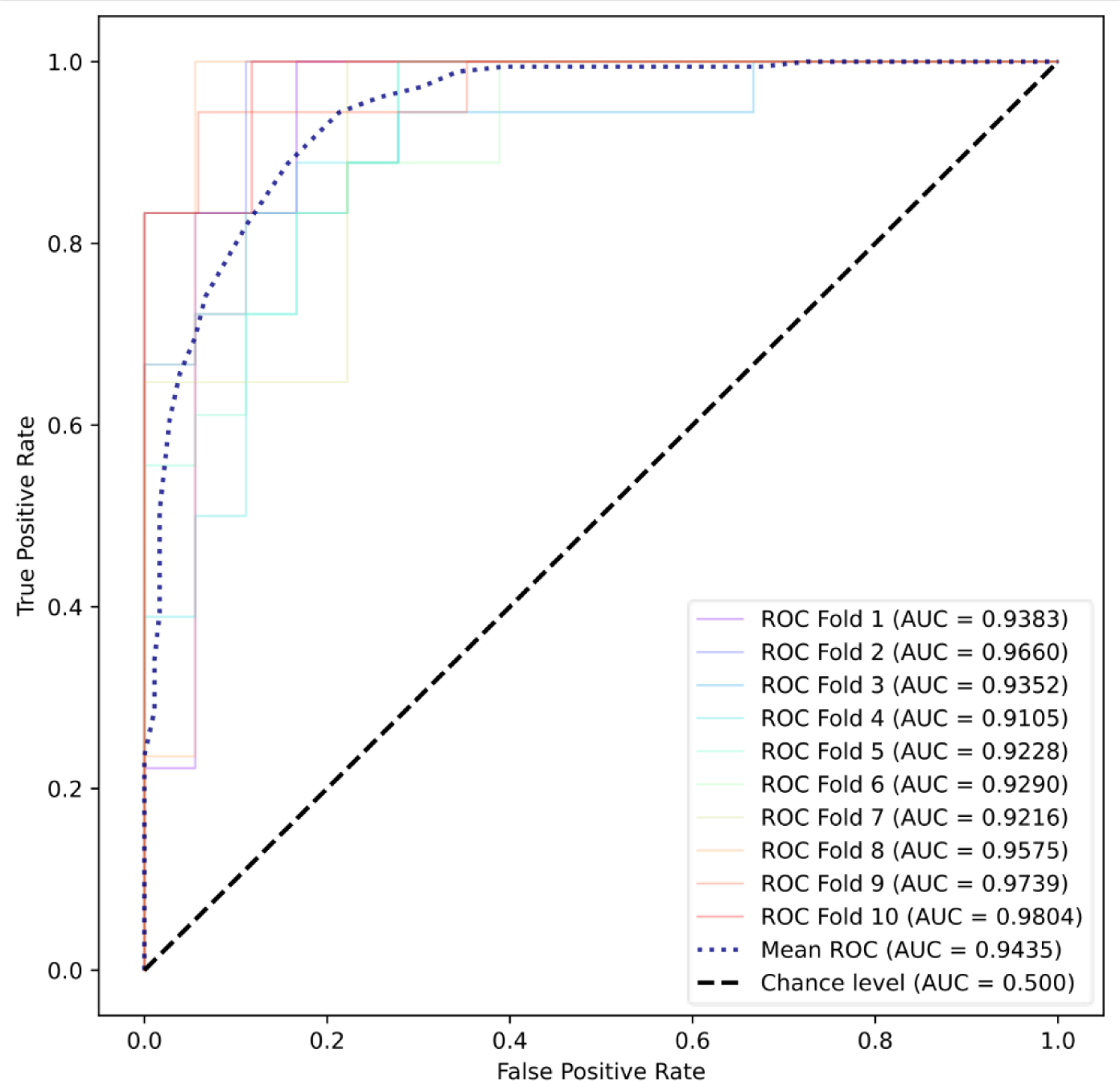
Binary classification WA 120_120 ROC curve based on frequency domain features.

**Table 3:**
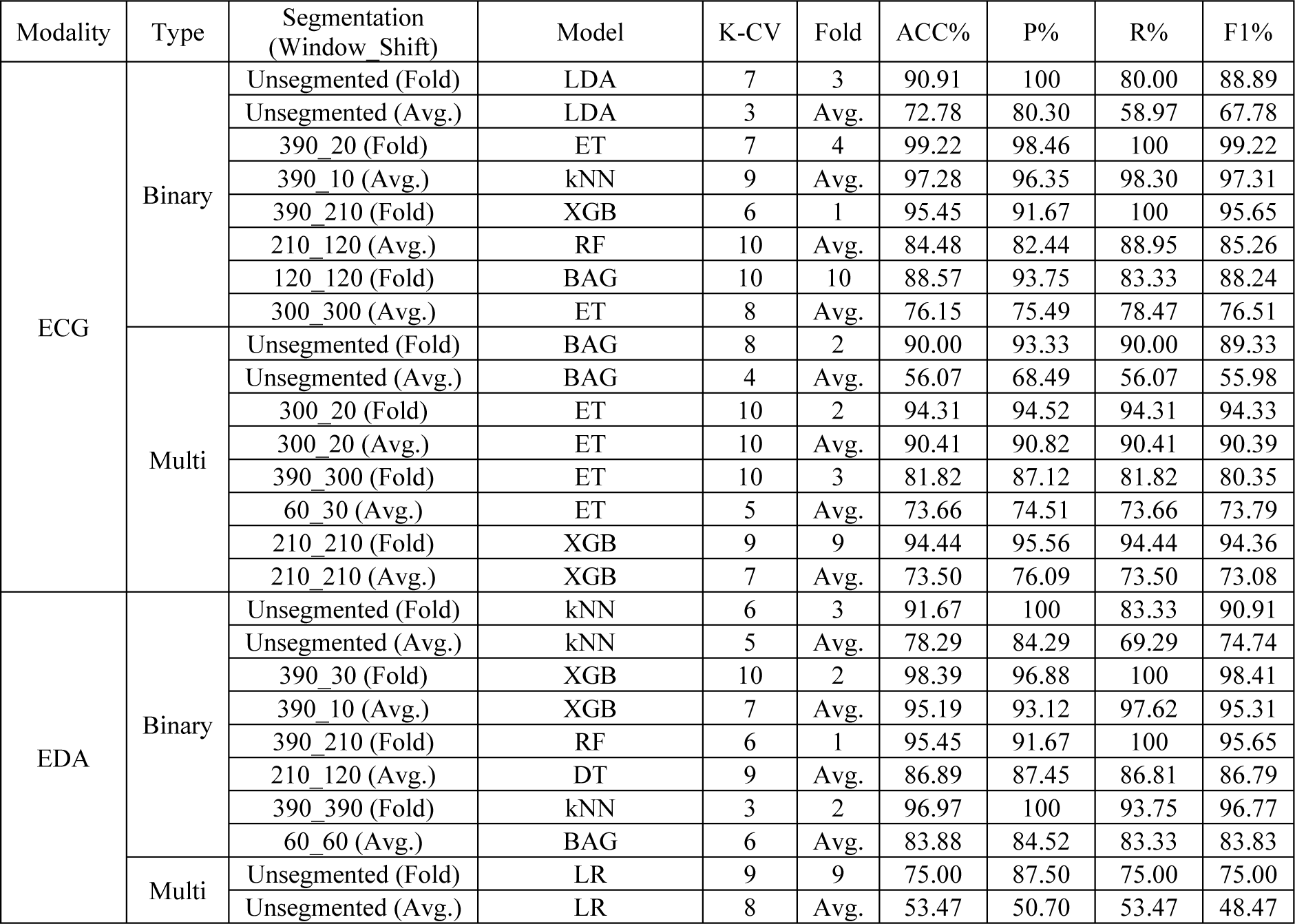
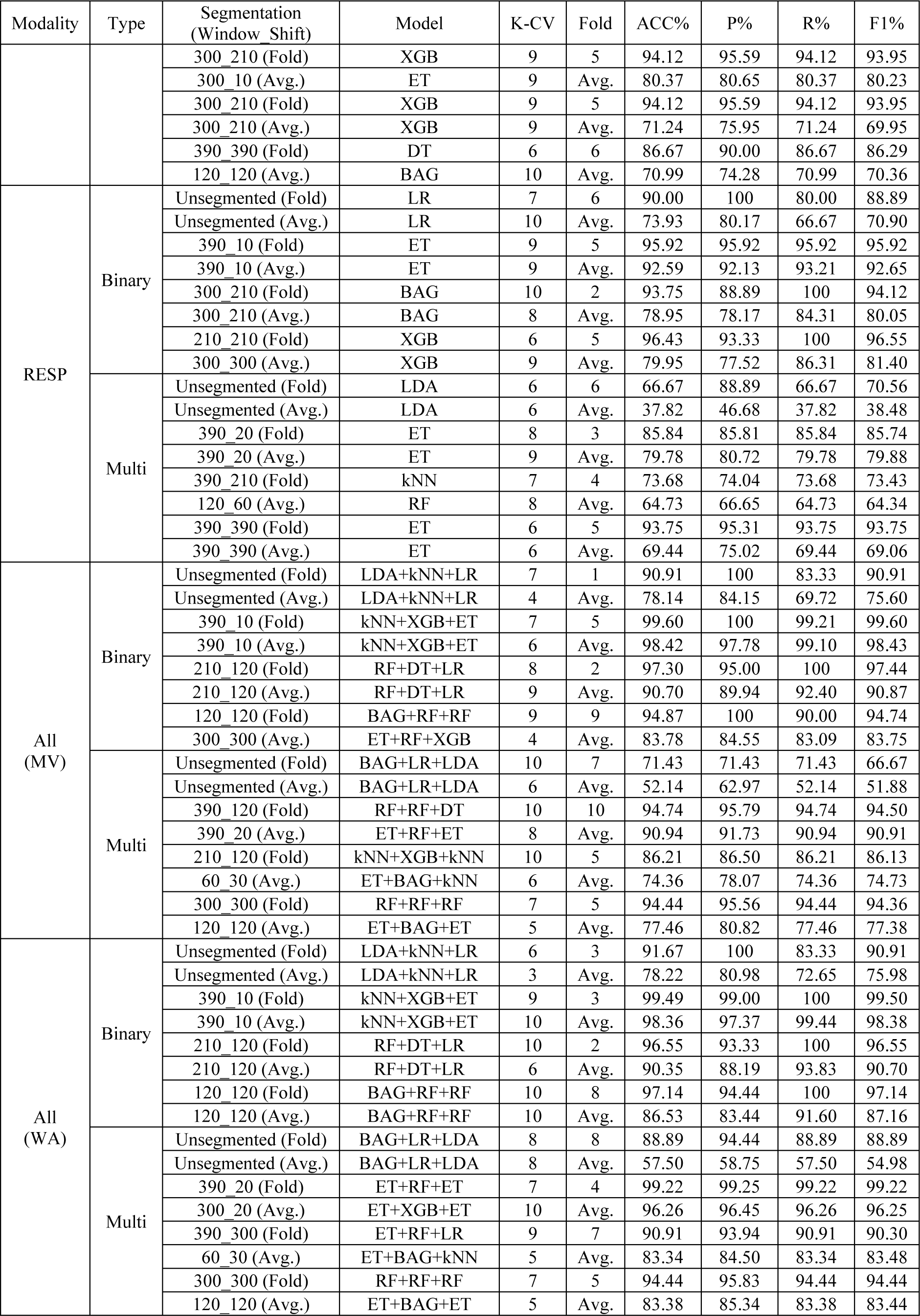
Best results based on frequency domain features.

| Modality | Type | Segmentation (Window_Shift) | Model | K-CV | Fold | ACC% | P% | R% | F1% |
| --- | --- | --- | --- | --- | --- | --- | --- | --- | --- |
| ECG | Binary | Unsegmented (Fold) | LDA | 7 | 3 | 90.91 | 100 | 80.00 | 88.89 |
|  |  | Unsegmented (Avg.) | LDA | 3 | Avg. | 72.78 | 80.30 | 58.97 | 67.78 |
|  |  | 390_20 (Fold) | ET | 7 | 4 | 99.22 | 98.46 | 100 | 99.22 |
|  |  | 390_10 (Avg.) | kNN | 9 | Avg. | 97.28 | 96.35 | 98.30 | 97.31 |
|  |  | 390_210 (Fold) | XGB | 6 | 1 | 95.45 | 91.67 | 100 | 95.65 |
|  |  | 210_120 (Avg.) | RF | 10 | Avg. | 84.48 | 82.44 | 88.95 | 85.26 |
|  |  | 120_120 (Fold) | BAG | 10 | 10 | 88.57 | 93.75 | 83.33 | 88.24 |
|  |  | 300_300 (Avg.) | ET | 8 | Avg. | 76.15 | 75.49 | 78.47 | 76.51 |
|  | Multi | Unsegmented (Fold) | BAG | 8 | 2 | 90.00 | 93.33 | 90.00 | 89.33 |
|  |  | Unsegmented (Avg.) | BAG | 4 | Avg. | 56.07 | 68.49 | 56.07 | 55.98 |
|  |  | 300_20 (Fold) | ET | 10 | 2 | 94.31 | 94.52 | 94.31 | 94.33 |
|  |  | 300_20 (Avg.) | ET | 10 | Avg. | 90.41 | 90.82 | 90.41 | 90.39 |
|  |  | 390_300 (Fold) | ET | 10 | 3 | 81.82 | 87.12 | 81.82 | 80.35 |
|  |  | 60_30 (Avg.) | ET | 5 | Avg. | 73.66 | 74.51 | 73.66 | 73.79 |
|  |  | 210_210 (Fold) | XGB | 9 | 9 | 94.44 | 95.56 | 94.44 | 94.36 |
|  |  | 210_210 (Avg.) | XGB | 7 | Avg. | 73.50 | 76.09 | 73.50 | 73.08 |
| EDA | Binary | Unsegmented (Fold) | kNN | 6 | 3 | 91.67 | 100 | 83.33 | 90.91 |
|  |  | Unsegmented (Avg.) | kNN | 5 | Avg. | 78.29 | 84.29 | 69.29 | 74.74 |
|  |  | 390_30 (Fold) | XGB | 10 | 2 | 98.39 | 96.88 | 100 | 98.41 |
|  |  | 390_10 (Avg.) | XGB | 7 | Avg. | 95.19 | 93.12 | 97.62 | 95.31 |
|  |  | 390_210 (Fold) | RF | 6 | 1 | 95.45 | 91.67 | 100 | 95.65 |
|  |  | 210_120 (Avg.) | DT | 9 | Avg. | 86.89 | 87.45 | 86.81 | 86.79 |
|  |  | 390_390 (Fold) | kNN | 3 | 2 | 96.97 | 100 | 93.75 | 96.77 |
|  |  | 60_60 (Avg.) | BAG | 6 | Avg. | 83.88 | 84.52 | 83.33 | 83.83 |
|  | Multi | Unsegmented (Fold) | LR | 9 | 9 | 75.00 | 87.50 | 75.00 | 75.00 |
|  |  | Unsegmented (Avg.) | LR | 8 | Avg. | 53.47 | 50.70 | 53.47 | 48.47 |

| Modality | Type | Segmentation<br>(Window_Shift) | Model | K-CV | Fold | ACC% | P% | R% | F1% |
| --- | --- | --- | --- | --- | --- | --- | --- | --- | --- |
|  |  | 300_210 (Fold) | XGB | 9 | 5 | 94.12 | 95.59 | 94.12 | 93.95 |
|  |  | 300_10 (Avg.) | ET | 9 | Avg. | 80.37 | 80.65 | 80.37 | 80.23 |
|  |  | 300_210 (Fold) | XGB | 9 | 5 | 94.12 | 95.59 | 94.12 | 93.95 |
|  |  | 300_210 (Avg.) | XGB | 9 | Avg. | 71.24 | 75.95 | 71.24 | 69.95 |
|  |  | 390_390 (Fold) | DT | 6 | 6 | 86.67 | 90.00 | 86.67 | 86.29 |
|  |  | 120_120 (Avg.) | BAG | 10 | Avg. | 70.99 | 74.28 | 70.99 | 70.36 |
| RESP | Binary | Unsegmented (Fold) | LR | 7 | 6 | 90.00 | 100 | 80.00 | 88.89 |
|  |  | Unsegmented (Avg.) | LR | 10 | Avg. | 73.93 | 80.17 | 66.67 | 70.90 |
|  |  | 390_10 (Fold) | ET | 9 | 5 | 95.92 | 95.92 | 95.92 | 95.92 |
|  |  | 390_10 (Avg.) | ET | 9 | Avg. | 92.59 | 92.13 | 93.21 | 92.65 |
|  |  | 300_210 (Fold) | BAG | 10 | 2 | 93.75 | 88.89 | 100 | 94.12 |
|  |  | 300_210 (Avg.) | BAG | 8 | Avg. | 78.95 | 78.17 | 84.31 | 80.05 |
|  |  | 210_210 (Fold) | XGB | 6 | 5 | 96.43 | 93.33 | 100 | 96.55 |
|  |  | 300_300 (Avg.) | XGB | 9 | Avg. | 79.95 | 77.52 | 86.31 | 81.40 |
|  | Multi | Unsegmented (Fold) | LDA | 6 | 6 | 66.67 | 88.89 | 66.67 | 70.56 |
|  |  | Unsegmented (Avg.) | LDA | 6 | Avg. | 37.82 | 46.68 | 37.82 | 38.48 |
|  |  | 390_20 (Fold) | ET | 8 | 3 | 85.84 | 85.81 | 85.84 | 85.74 |
|  |  | 390_20 (Avg.) | ET | 9 | Avg. | 79.78 | 80.72 | 79.78 | 79.88 |
|  |  | 390_210 (Fold) | kNN | 7 | 4 | 73.68 | 74.04 | 73.68 | 73.43 |
|  |  | 120_60 (Avg.) | RF | 8 | Avg. | 64.73 | 66.65 | 64.73 | 64.34 |
|  |  | 390_390 (Fold) | ET | 6 | 5 | 93.75 | 95.31 | 93.75 | 93.75 |
|  |  | 390_390 (Avg.) | ET | 6 | Avg. | 69.44 | 75.02 | 69.44 | 69.06 |
| All<br>(MV) | Binary | Unsegmented (Fold) | LDA+kNN+LR | 7 | 1 | 90.91 | 100 | 83.33 | 90.91 |
|  |  | Unsegmented (Avg.) | LDA+kNN+LR | 4 | Avg. | 78.14 | 84.15 | 69.72 | 75.60 |
|  |  | 390_10 (Fold) | kNN+XGB+ET | 7 | 5 | 99.60 | 100 | 99.21 | 99.60 |
|  |  | 390_10 (Avg.) | kNN+XGB+ET | 6 | Avg. | 98.42 | 97.78 | 99.10 | 98.43 |
|  |  | 210_120 (Fold) | RF+DT+LR | 8 | 2 | 97.30 | 95.00 | 100 | 97.44 |
|  |  | 210_120 (Avg.) | RF+DT+LR | 9 | Avg. | 90.70 | 89.94 | 92.40 | 90.87 |
|  |  | 120_120 (Fold) | BAG+RF+RF | 9 | 9 | 94.87 | 100 | 90.00 | 94.74 |
|  |  | 300_300 (Avg.) | ET+RF+XGB | 4 | Avg. | 83.78 | 84.55 | 83.09 | 83.75 |
|  | Multi | Unsegmented (Fold) | BAG+LR+LDA | 10 | 7 | 71.43 | 71.43 | 71.43 | 66.67 |
|  |  | Unsegmented (Avg.) | BAG+LR+LDA | 6 | Avg. | 52.14 | 62.97 | 52.14 | 51.88 |
|  |  | 390_120 (Fold) | RF+RF+DT | 10 | 10 | 94.74 | 95.79 | 94.74 | 94.50 |
|  |  | 390_20 (Avg.) | ET+RF+ET | 8 | Avg. | 90.94 | 91.73 | 90.94 | 90.91 |
|  |  | 210_120 (Fold) | kNN+XGB+kNN | 10 | 5 | 86.21 | 86.50 | 86.21 | 86.13 |
|  |  | 60_30 (Avg.) | ET+BAG+kNN | 6 | Avg. | 74.36 | 78.07 | 74.36 | 74.73 |
|  |  | 300_300 (Fold) | RF+RF+RF | 7 | 5 | 94.44 | 95.56 | 94.44 | 94.36 |
|  |  | 120_120 (Avg.) | ET+BAG+ET | 5 | Avg. | 77.46 | 80.82 | 77.46 | 77.38 |
| All<br>(WA) | Binary | Unsegmented (Fold) | LDA+kNN+LR | 6 | 3 | 91.67 | 100 | 83.33 | 90.91 |
|  |  | Unsegmented (Avg.) | LDA+kNN+LR | 3 | Avg. | 78.22 | 80.98 | 72.65 | 75.98 |
|  |  | 390_10 (Fold) | kNN+XGB+ET | 9 | 3 | 99.49 | 99.00 | 100 | 99.50 |
|  |  | 390_10 (Avg.) | kNN+XGB+ET | 10 | Avg. | 98.36 | 97.37 | 99.44 | 98.38 |
|  |  | 210_120 (Fold) | RF+DT+LR | 10 | 2 | 96.55 | 93.33 | 100 | 96.55 |
|  |  | 210_120 (Avg.) | RF+DT+LR | 6 | Avg. | 90.35 | 88.19 | 93.83 | 90.70 |
|  |  | 120_120 (Fold) | BAG+RF+RF | 10 | 8 | 97.14 | 94.44 | 100 | 97.14 |
|  |  | 120_120 (Avg.) | BAG+RF+RF | 10 | Avg. | 86.53 | 83.44 | 91.60 | 87.16 |
|  | Multi | Unsegmented (Fold) | BAG+LR+LDA | 8 | 8 | 88.89 | 94.44 | 88.89 | 88.89 |
|  |  | Unsegmented (Avg.) | BAG+LR+LDA | 8 | Avg. | 57.50 | 58.75 | 57.50 | 54.98 |
|  |  | 390_20 (Fold) | ET+RF+ET | 7 | 4 | 99.22 | 99.25 | 99.22 | 99.22 |
|  |  | 300_20 (Avg.) | ET+XGB+ET | 10 | Avg. | 96.26 | 96.45 | 96.26 | 96.25 |
|  |  | 390_300 (Fold) | ET+RF+LR | 9 | 7 | 90.91 | 93.94 | 90.91 | 90.30 |
|  |  | 60_30 (Avg.) | ET+BAG+kNN | 5 | Avg. | 83.34 | 84.50 | 83.34 | 83.48 |
|  |  | 300_300 (Fold) | RF+RF+RF | 7 | 5 | 94.44 | 95.83 | 94.44 | 94.44 |
|  |  | 120_120 (Avg.) | ET+BAG+ET | 5 | Avg. | 83.38 | 85.34 | 83.38 | 83.44 |

#### 4.2.1 Performance Evaluation of ECG Modality

As presented in Table 3, the performance of the ECG modality was assessed across different segmentation strategies and classifiers for binary classification. With unsegmented ECG data, the optimized LDA classifier achieved the highest accuracy of 72.78% using 3-CV. For segmentations with over 50% overlap, a window of 390 seconds with a 10-second shift yielded the highest accuracy of 97.28% using an optimized kNN classifier and 9-CV. In cases of segmentation with 50% and under overlap, a window of 210 seconds with a 120-second shift resulted in the highest accuracy of 84.48% with an optimized RF classifier and 10-CV. For segmentation with no overlap, a window of 300 seconds provided the highest accuracy of 76.15% using an optimized ET classifier and 8-CV. These results are illustrated in Fig. 5, providing a visual summary of the performance across different segmentation strategies.

As detailed in Table 3, the evaluation of the ECG modality for multi-class classification revealed varying performance across different segmentation approaches and classifiers. With unsegmented ECG data, the optimized BAG classifier achieved the highest accuracy of 56.07% using 4-CV. For segmentations with over 50% overlap, a window of 300 seconds with a 20-second shift yielded the highest accuracy of 90.41% using an optimized ET classifier and 10-CV. In cases of segmentation with 50% and under overlap, a window of 60 seconds with a 30-second shift resulted in the highest accuracy of 73.66% with an optimized ET classifier and 5-CV. For segmentation with no overlap, a window of 210 seconds provided the highest accuracy of 73.50% using an optimized XGB classifier and 7-CV. Fig. 6 visually summarizes these results, highlighting the performance variations across different segmentation strategies.

#### 4.2.2 Performance Evaluation of EDA Modality

As presented in Table 3, the performance of the EDA modality was assessed across different segmentation strategies and classifiers for binary classification. With unsegmented EDA data, the optimized kNN classifier achieved the highest accuracy of 78.29% using 5-CV. For segmentations with over 50% overlap, a window of 390 seconds with a 10-second shift yielded the highest accuracy of 95.19% using an optimized XGB classifier and 7-CV. In cases of segmentation with 50% and under overlap, a window of 210 seconds with a 120-second shift resulted in the highest accuracy of 86.89% with an optimized DT classifier and 9-CV. For segmentation with no overlap, a window of 60 seconds provided the highest accuracy of 83.88% using an optimized BAG classifier and 6-CV. These results are illustrated in Fig. 5, providing a visual summary of the performance across different segmentation strategies.

As detailed in Table 3, the evaluation of the EDA modality for multi-class classification revealed varying performance across different segmentation approaches and classifiers. With unsegmented EDA data, the optimized LR classifier achieved the highest accuracy of 53.47% using 8-CV. For segmentations with over 50% overlap, a window of 300 seconds with a 10-second shift yielded the highest accuracy of 80.37% using an optimized ET classifier and 9-CV. In cases of segmentation with 50% and under overlap, a window of 300 seconds with a 210-second shift resulted in the highest accuracy of 71.24% with an optimized XGB classifier and 9-CV. For segmentation with no overlap, a window of 120 seconds provided the highest accuracy of 70.99% using an optimized BAG classifier and 10-CV. Fig. 6 visually summarizes these results, highlighting the performance variations across different segmentation strategies.

#### 4.2.3 Performance Evaluation of RESP Modality

As presented in Table 3, the performance of the RESP modality was assessed across different segmentation strategies and classifiers for binary classification. With unsegmented RESP data, the optimized LR classifier achieved the highest accuracy of 73.93% using 10-CV. For segmentations with over 50% overlap, a window of 390 seconds with a 10-second shift yielded the highest accuracy of 92.59% using an optimized ET classifier and 9-CV. In cases of segmentation with 50% and under overlap, a window of 300 seconds with a 210-second shift resulted in the highest accuracy of 78.95% with an optimized BAG classifier and 8-CV. For segmentation with no overlap, a window of 300 seconds provided the highest accuracy of 79.95% using an optimized XGB classifier and 9-CV. These results are illustrated in Fig. 5, providing a visual summary of the performance across different segmentation strategies.

As detailed in Table 3, the evaluation of the RESP modality for multi-class classification revealed varying performance across different segmentation approaches and classifiers. With unsegmented RESP data, the optimized LDA classifier achieved the highest accuracy of 37.82% using 6-CV. For segmentations with over 50% overlap, a window of 390 seconds with a 20-second shift yielded the highest accuracy of 79.78% using an optimized ET classifier and 9-CV. In cases of segmentation with 50% and under overlap, a window of 120 seconds with a 60-second shift resulted in the highest accuracy of 64.73% with an optimized RF classifier and 8-CV. For segmentation with no overlap, a window of 390 seconds provided the highest accuracy of 69.44% using an optimized ET classifier and 6-CV. Fig. 6 visually summarizes these results, highlighting the performance variations across different segmentation strategies.

#### 4.2.4 Performance Evaluation of Modalities MV Ensemble Method

As presented in Table 3, the performance of the modalities MV ensemble was assessed across different segmentation strategies and classifiers for binary classification. With unsegmented MV Ensemble data, the optimized LDA, kNN, and LR classifiers for ECG, EDA, and RESP respectively achieved the highest accuracy of 78.14% using 4-CV. For segmentations with over 50% overlap, a window of 390 seconds with a 10-second shift yielded the highest accuracy of 98.42% using optimized kNN, XGB, and ET classifiers for ECG, EDA, and RESP respectively, and 6-CV. In cases of segmentation with 50% and under overlap, a window of 210 seconds with a 120-second shift resulted in the highest accuracy of 90.70% with optimized RF, DT, and LR classifiers for ECG, EDA, and RESP respectively, and 9-CV. For segmentation with no overlap, a window of 300 seconds provided the highest accuracy of 83.78% using optimized ET, RF, and XGB classifiers for ECG, EDA, and RESP respectively, and 4-CV. These results are illustrated in Fig. 5, providing a visual summary of the performance across different segmentation strategies.

As detailed in Table 3, the evaluation of the modalities MV ensemble for multi-class classification revealed varying performance across different segmentation approaches and classifiers. With unsegmented MV Ensemble data, the optimized BAG, LR, and LDA classifiers for ECG, EDA, and RESP respectively achieved the highest accuracy of 52.14% using 6-CV. For segmentations with over 50% overlap, a window of 390 seconds with a 20-second shift yielded the highest accuracy of 90.94% using optimized ET, RF, and ET classifiers for ECG, EDA, and RESP respectively, and 8-CV. In cases of segmentation with 50% and under overlap, a window of 60 seconds with a 30-second shift resulted in the highest accuracy of 74.36% with optimized ET, BAG, and kNN classifiers for ECG, EDA, and RESP respectively, and 6-CV. For segmentation with no overlap, a window of 120 seconds provided the highest accuracy of 77.46% using optimized ET, BAG, and ET classifiers for ECG, EDA, and RESP respectively, and 5-CV. Fig. 6 visually summarizes these results, highlighting the performance variations across different segmentation strategies.

#### 4.2.5 Performance Evaluation of Modalities WA Ensemble Method

As presented in Table 3, the performance of the modalities WA ensemble was assessed across different segmentation strategies and classifiers for binary classification. With unsegmented WA Ensemble data, the optimized LDA, kNN, and LR classifiers for ECG, EDA, and RESP respectively achieved the highest accuracy of 78.22% using 3-CV. For segmentations with over 50% overlap, a window of 390 seconds with a 10-second shift yielded the highest accuracy of 98.36% using optimized kNN, XGB, and ET classifiers for ECG, EDA, and RESP respectively, and 10-CV. In cases of segmentation with 50% and under overlap, a window of 210 seconds with a 120-second shift resulted in the highest accuracy of 90.35% with optimized RF, DT, and LR classifiers for ECG, EDA, and RESP respectively, and 6-CV. For segmentation with no overlap, a window of 120 seconds provided the highest accuracy of 86.53% using optimized BAG, RF, and RF classifiers for ECG, EDA, and RESP respectively and 10-CV, additionally ROC is depicted in Fig. 7. These results are illustrated in Fig. 5, providing a visual summary of the performance across different segmentation strategies.

As detailed in Table 3, the evaluation of the modalities WA ensemble for multi-class classification revealed varying performance across different segmentation approaches and classifiers. With unsegmented WA Ensemble data, the optimized BAG, LR, and LDA classifiers for ECG, EDA, and RESP respectively achieved the highest accuracy of 57.50% using 8-CV. For segmentations with over 50% overlap, a window of 300 seconds with a 20-second shift yielded the highest accuracy of 96.26% using optimized ET, XGB, and ET classifiers for ECG, EDA, and RESP respectively, and 10-CV. In cases of segmentation with 50% and under overlap, a window of 60 seconds with a 30-second shift resulted in the highest accuracy of 83.34% with optimized ET, BAG, and kNN classifiers for ECG, EDA, and RESP respectively, and 5-CV. For segmentation with no overlap, a window of 120 seconds provided the highest accuracy of 83.38% using optimized ET, BAG, and ET classifiers for ECG, EDA, and RESP respectively, and 5-CV. Fig. 6 visually summarizes these results, highlighting the performance variations across different segmentation strategies.

## 5. Discussion

In this study, we investigated the impact of wearable devices on the early detection of psychological stress, employing both binary and five-class classifications. Our findings reveal significant correlations between the modalities, ECG, EDA, and RESP, and stress levels, suggesting the efficacy of these biomarkers for stress detection. Additionally, we employed two ensemble methods that simultaneously integrated these modalities. These results are consistent with previous research (e.g., [14], [16], [17], [19], [20], [21], [23], [25]), which also highlighted the utility of physiological signals in stress monitoring. Our investigation into five-class stress classification and the use of ensemble methods provides a novel contribution to the field. These results suggest that wearable devices could significantly enhance stress monitoring and management, improving overall quality of life. Future research should aim to address current limitations and refine these models further.

### 5.1. Comparative Evaluation of Methodologies

Table 4 juxtaposes our research findings with relevant studies, delineating comparisons based on the utilization of individual modalities, namely, ECG, EDA, and RESP. Each of our employed modalities is juxtaposed against corresponding studies employing singular modalities. Additionally, our ensemble methodology is compared against studies integrating all three modalities simultaneously.

**Table 4:** Comparison with related studies.

| Paper References | Dataset | Modality | Validation | Model | ACC% | Our Proposal |
| --- | --- | --- | --- | --- | --- | --- |
| Zhu et al (2023) [32] | CLAS | EDA<br>Non-overlap | LOSO | SVM | 68.5 | EDA<br>120_120<br>9-CV<br>kNN<br>**86.79** |
|  | UTD |  |  | RF | 73.1 |  |
|  | VerBIO |  |  | SVM | 92.9 |  |
|  | WESAD |  |  | RF | **86.5** |  |
| Adarsh et al (2024) [33] | WESAD | ECG<br>Overlap>50% | 5-CV | GCN | **97.75** | ECG<br>300_10<br>8-CV<br>BAG<br>**99.63** |
|  | SWELL |  |  |  | 94.48 |  |
| Schmidt et al (2018) [16] | WESAD | RESP<br>Overlap>50% | LOSO | LDA | 88.09 | RESP<br>300_10<br>9-CV<br>XGB<br>**99.43** |
| Rashid et al (2023) [34] | WESAD | ECG+EDA+RESP<br>Overlap>50% | LOSO | AB | **81.62** | WA<br>300_10<br>4-CV<br>BAG+XGB+<br>kNN<br>**99.96** |
|  |  | ACCE+ECG+EDA<br>Overlap>50% |  |  | 86.37 |  |

Zhu et al (2023) [32] conducted research on binary stress classification utilizing exclusively the EDA modality from four distinct datasets: CLAS, UTD, VerBIO, and WESAD. The primary focus is on the WESAD dataset, which our study employed. In their investigation, an accuracy of 86.5% was achieved by utilizing segmentation without overlap, employing a 30-second window size, and employing the RF classifier with LOSO cross-validation. Conversely, our study attained a slightly higher accuracy of 86.79% under similar settings, exclusively utilizing the EDA modality, employing segmentation without overlap, employing a 120-second window size, and utilizing the kNN classifier with 9-CV.

Adarsh et al (2024) [33] investigated binary stress classification by leveraging the ECG modality from two distinct datasets: SWELL and WESAD. The primary focus is on the WESAD dataset, which our study employed. In the research conducted by the authors, an accuracy of 97.75% was achieved through segmentation with more than 50% overlap, utilizing a window size of 5 seconds with a 0.25-second shift, and employing graph convolutional Networks (GCN) with 5-CV. Conversely, the investigation conducted in our study yielded a higher accuracy of 99.63% under analogous conditions, exclusively employing the ECG modality, employing segmentation with more than 50% overlap, utilizing a window size of 300 seconds with a 10-second shift, and employing the BAG classifier with 8-CV.

Schmidt et al (2018) [16] explored binary stress classification by utilizing the Respiration (RESP) modality from the WESAD dataset, which was also employed in our study. In the investigation conducted by Schmidt et al, an accuracy of 88.09% was achieved through segmentation with more than 50% overlap, employing a window size of 60 seconds with a 0.25-second shift, and utilizing LDA with LOSO cross-validation. Conversely, our study attained a higher accuracy of 99.43% under similar conditions, exclusively utilizing the RESP modality, employing segmentation with more than 50% overlap, utilizing a window size of 300 seconds with a 10-second shift, and employing the XGB classifier with 9-CV.

Rashid et al (2023) [34] investigated binary stress classification through a multimodal approach incorporating ECG, EDA, and RESP modalities from the WESAD dataset, which aligns with the dataset utilized in our study. In their investigation, Rashid et al. achieved an accuracy of 81.62% by employing segmentation with more than 50% overlap, utilizing a window size of 60 seconds with a 5-second shift, and employing AB with LOSO cross-validation. Conversely, our study achieved a notably higher accuracy of 99.96% under analogous conditions, employing a multimodal WA ensemble of ECG, EDA, and RESP modalities. Our methodology involved segmentation with more than 50% overlap, utilizing a window size of 300 seconds with a 10-second shift, and employing BAG, XGB, and kNN classifiers for ECG, EDA, and RESP, respectively, with 4-CV.

### 5.2. Principle Findings

We utilized three different modalities: ECG, EDA, and RESP. Additionally, we employed two ensemble methods that simultaneously integrated these modalities. Our investigation into commercially available wearable devices that offer these modalities is summarized in Table 5, derived from Taskasaplidis et al (2024) [35].

**Table 5:** Commercial wearable devices that provide ECG, EDA, and RESP modalities.

| Device Name | Body Location | ECG | EDA | RESP | Additional Features |
| --- | --- | --- | --- | --- | --- |
| Fitbit Sense 2 | Wrist | Yes | Yes | Yes | SpO2, SKT |
| Flowtime | Head | Yes | No | No | 2-channel brainwave |
| Movesense | Chest | Yes | No | No | Motion measurement |
| Prana | Weist | No | No | Yes | Posture |
| Sentio Solutions Feel Therapeutics | Wrist | Yes | Yes | No | Physical activity, SKT |

Based on our findings, we recommend the configurations presented in Table 6 for optimal performance in real-world applications tailored to the system type and available modalities.

**Table 6:**
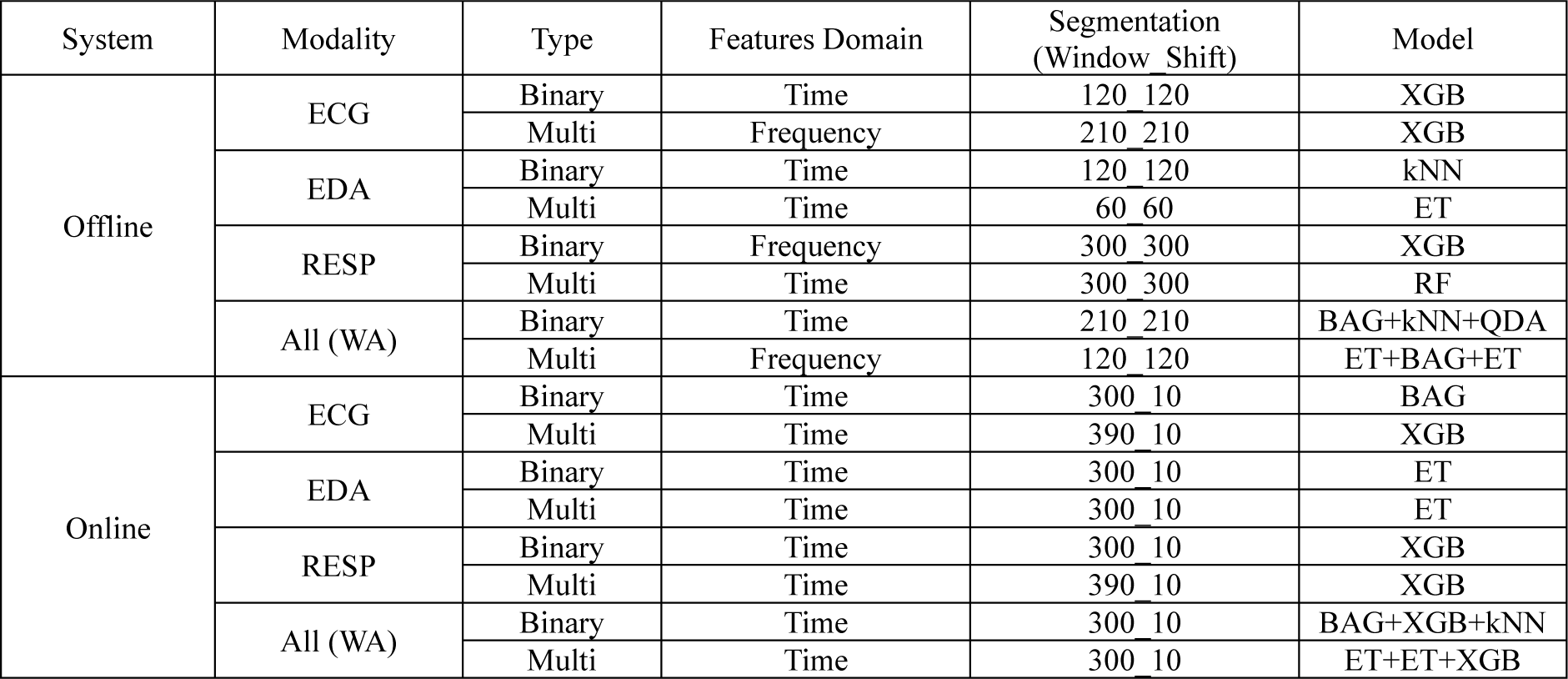
Recommended Configurations for Optimal Performance.

## 6. Conclusion

This study explored the effectiveness of wearable devices in the early detection of psychological stress, utilizing both binary and five-class classification models. Our findings demonstrated significant correlations between stress levels and physiological signals from ECG, EDA, and RESP, confirming these modalities as reliable biomarkers for stress detection. We tested ten different classifiers: Random Forest (RF), Extreme Gradient Boosting (XGB), k-nearest Neighbors (kNN), Logistic Regression (LR), Decision Tree (DT), AdaBoost (AB), Extra Trees (ET), Bagging (BAG), Quadratic Discriminant Analysis (QDA), and Linear Discriminant Analysis (LDA). We also applied hyperparameter optimization using grid search, incorporating time and frequency domain features separately in our analyses. We employed two ensemble methods, Majority Voting (MV) and Weighted Averaging (WA), to integrate these modalities, enhancing the accuracy and robustness of the stress detection system. Additionally, we reviewed commercially available wearable devices capable of providing these physiological measurements. Based on our findings, we recommend the configurations detailed in Table 6 for optimal performance in real-world applications. These recommendations are tailored to the specific system types and available modalities, ensuring maximum effectiveness and utility. This research underscores the potential of multimodal wearable devices in the early detection and monitoring of psychological stress, offering a foundation for future research and practical applications in wearable health technology. While our study provides valuable insights, future research could benefit from exploring the integration of both time and frequency domain features, as well as investigating the potential of deep learning models to enhance detection capabilities further.

## Data availability and materials

The data used in the publication is publicly available.

## Code availability

The developed code can be provided by the Corresponding Author upon reasonable request.

## Ethical approval

No ethical approval is required for this study.

## Author contributions

**Basil A. Darwish:** Writing – original draft, Methodology, Formal analysis, Data curation. **Nancy M. Salem:** Writing – review & editing, Validation, Supervision, Methodology, Formal analysis, Conceptualization. **Ghada Kareem:** Writing – review & editing, Validation, Methodology, Conceptualization. **Lamees N. Mahmoud:** Writing – review & editing, Validation, Methodology, Conceptualization. **Ibrahim Sadek:** Writing – review & editing, Validation, Supervision, Methodology, Formal analysis, Conceptualization.

## Appendix: List of Abbreviations

AB: AdaBoost
ACC: Accuracy
ACCE: Three-Axis Acceleration
AI: Artificial Intelligence
Avg: Average
BAG: Bagging
BVP: Blood Volume Pulse
cGAN: Conditional Generative Adversarial Network
CNN: Convolutional Neural Network
DL: Deep Learning
DNN: Deep Neural Network
DT: Decision Tree
ECG: Electrocardiogram
EDA: Electrodermal Activity
ET: Extra Trees
F1: F1 Score
FCN: Fully Convolutional Network
FFT: Fast Fourier Transform
GAN: Generative Adversarial Network
GCN: Graph Convolutional Networks
GSR: Galvanic Skin Response
HR: Heart Rate
HRV: Heart Rate Variability
IMU: Inertial Measurement Unit
kNN: k-Nearest Neighbors
K-CV: K-fold Cross-Validation
LDA: Linear Discriminant Analysis
LOSO: Leave-One-Subject-Out
LR: Logistic Regression
LSTM: Long Short-Term Memory
ML: Machine Learning
MLP: Multi-Layer Perceptron
MMTM: Multimodal Transfer Module
MV: Majority Voting
P: Precision
PANAS: Positive and Negative Affect Schedule Questionnaire
PPG: Photoplethysmography
PRV: Pulse Rate Variability
PSD: Power Spectral Density
QDA: Quadratic Discriminant Analysis
R: Recall
RESP: Respiration
RF: Random Forest
ROC: Receiver Operating Characteristic
SFM: Select From Model
SKT: Skin Temperature
SVM: Support Vector Machine
TEMP: Body Temperature
TSST: Trier Social Stress Test
WA: Weighted Average
WESAD: Wearable Stress and Affect Detection Dataset
XGB: Extreme Gradient Boosting

